# Distinct and shared genetics of kidney filtration function versus albuminuria revealed by multi-trait GWAS

**DOI:** 10.64898/2026.06.08.26355141

**Authors:** Hannah C. de Hesselle, B. Frederike Garben, Klaus J. Stark, Richard Warth, Alexander Teumer, Cristian Pattaro, Iris M. Heid, Thomas W Winkler

## Abstract

Chronic kidney disease is characterized by decreased glomerular filtration rate (eGFR, estimated from serum creatinine or cystatin C) or increased urinary albumin-to-creatinine-ratio (UACR). Genome-wide association studies provided the genetic make-up of these traits, but their overlap remained largely unknown. Our multi-trait GWAS (N=1M) identified 812 signals and multi-trait fine-mapping sharpened the identification of likely causal variants. Of 333 signals classified for filtration function or albuminuria, only 11 overlapped. Their effects on eGFR and UACR were directionally concordant, dominated by eGFR and independent of HbA1c or mean arterial pressure. Mapped genes pinpointed mechanisms related to glomerular filtration area (*SHROOM3*, *EPB41L5*) and sodium-mediated intraglomerular pressure (*NRBP1*, *DPEP1*/*CHMP1A*). Genetics of fluid intake resulted in shadow effects on UACR without albumin leakage into urine. Our multi-trait approach sharpened the identification of likely causal genes for kidney traits, demonstrated largely distinct genetics for filtration function versus albuminuria, and provided new biological insights into the overlap.

## INTRODUCTION

Chronic kidney disease (CKD) is a common, severe disease. It affects about 10% of the adults globally^1,2^ and affected individuals are at substantially increased risk for kidney failure and cardiovascular mortality^3,4^.

The definition of CKD is multi-dimensional. Two components of CKD are abnormalities in kidney function or kidney structure^5^. Kidney function is reflected by the glomerular filtration rate (GFR), which is typically estimated based on serum creatinine (eGFRcrea) or cystatin C (eGFRcys). Another marker of kidney function is blood urea nitrogen^6^ (BUN). A marker of structural kidney damage is albuminuria, assessed by the urinary albumin-to-creatinine ratio (UACR). CKD is classified into higher stages by decreased levels of GFR (G1-G5) and increased degrees of albuminuria (A1-A3) by a risk stratification matrix provided by the “Kidney Disease Improving Global Outcomes” organization^5^ (KDIGO matrix). The KDIGO stages characterize the risk for kidney failure, which increases by decreased levels of GFR and increased degree of albuminuria jointly, but also independently. Unraveling the genetics basis of CKD by integrating both filtration function and albuminuria can help understand shared and distinct mechanisms. This can be pivotal to improve diagnosis, prevention, and treatment strategies.

Over the past decades, genome-wide association studies (GWAS) have significantly advanced our understanding of kidney trait genetics. GWAS have uncovered hundreds of loci linked to eGFR^7–10^ or UACR^11^. However, these studies focused on single-trait GWAS, and a joint view and systematic comparison of eGFR and UACR genetics has been limited so far. The only comparison of genetic effects on eGFR and UACR, from 2012, was limited to 16 candidate variants^12^. Only one variant, located in the *SHROOM3* gene, was found with significant association for both eGFR and UACR based on 31,580 individuals of the general adult population. The eGFR-lowering allele was associated with lower UACR, which was an unexpected directionality given the known increased risk of kidney failure by lower eGFR and higher UACR. This was attributed to independent effects on both traits (pleiotropy) or higher eGFR linked to albuminuria in hyperfiltration states^12^. The question remained whether the *SHROOM3* variant is a singular phenomenon and the genetics of eGFR and UACR in the general population is truly distinct or further overlap was missed due to limited power. Importantly, when individual participants data is available at large enough sample size, mediator analyses can dissect independent from indirect effects of eGFR and UACR and help understand the link to potentially linked traits like perfusion pressure or diabetes.

Recently, multi-trait methods for GWAS and fine-mapping have emerged, offering enhanced power to detect joint associations and improved precision to identify causal variants by leveraging the genetic correlation across related traits^13–16^. Despite the availability of these methods, there is a notable lack of GWAS integrating multiple kidney traits. This results in missed opportunities to sharpen association signals and understand similarities and differences between eGFR and UACR genetics and underlying pathways.

Therefore, we aimed to refine our understanding of the genetic architecture of CKD regarding shared and distinct mechanisms that modulate GFR versus UACR and to help prioritize novel causal genes. We approached this by (i) a large multi-trait GWAS and independent signal detection using GWAS summary statistics of eGFRcrea, eGFRcys, BUN, and UACR from UK Biobank (UKB) and the Chronic Kidney Disease Genetics Consortium (CKDGen Consortium, N up to 1 million) and (ii) multi-trait fine-mapping to map credibly causal variants to likely causal genes. For independent signal detection, we developed a new sample size unbiased approach. We (iii) ascertained genetic variant relevance for “real eGFR” opposed to mere biomarker metabolism by joint effects on eGFRcrea and eGFRcys and relevance for “real UACR” opposed to mere urinary creatinine (Ucrea) associations by joint effects on UACR and urinary albumin (Ualb). Finally, we (iv) classified identified signals as “real eGFR only”, “real UACR only”, or “both” using UKB (N up to 450,000) followed by biological annotation and mediator analyses to help understand mechanisms (overview in **Supplementary Figure 1**).

## RESULTS

### Multi-trait GWAS meta-analysis identifies 812 kidney trait signals

First, we established the broad basis of genetics of kidney function traits and evaluated whether a multi-trait GWAS approach improved power to identify loci and independent signals. For this, we used European-ancestry single-trait GWAS summary statistics for four traits based on meta-analyses of CKDGen and UKB (**Supplementary Table 1, Methods**): eGFRcrea^8^ (N=1,004,040), eGFRcys^8^ (N=460,826), BUN (N= 679,532; newly generated) and UACR^11^ (N=547,361). We obtained significant genomic regions from single-trait GWAS across the four traits or multi-trait GWAS (C-GWAS^14^, **Methods**). We identified 575 distinct genomic regions with genome-wide significant associations (P<5×10^-8^ for at least one trait, or P_C-GWAS_<5×10^-8^, >500 kb distance between regions, **Figure 1A**, **Supplementary Figure 2, Supplementary Table 2, Methods**). Among them, 23 were only identified with genome-wide significance by C-GWAS and not by any of the four single traits, including 9 completely novel kidney trait loci compared to previous work^8–11^ (**Supplementary Figure 3, Supplementary Table 2**). For example, signals near *LOC440040* and *ZFHX4*, both known to be implicated in renal cell carcinoma, were only identified by C-GWAS^17,18^. C-GWAS alone captured most, 451, of the 575 regions; many loci were identified via eGFRcrea due to the large sample size of this trait (**Figure 1B**). C-GWAS P-values tended to be smaller for variants implicated by more than two traits (**Figure 1C**).

**Figure 1:**
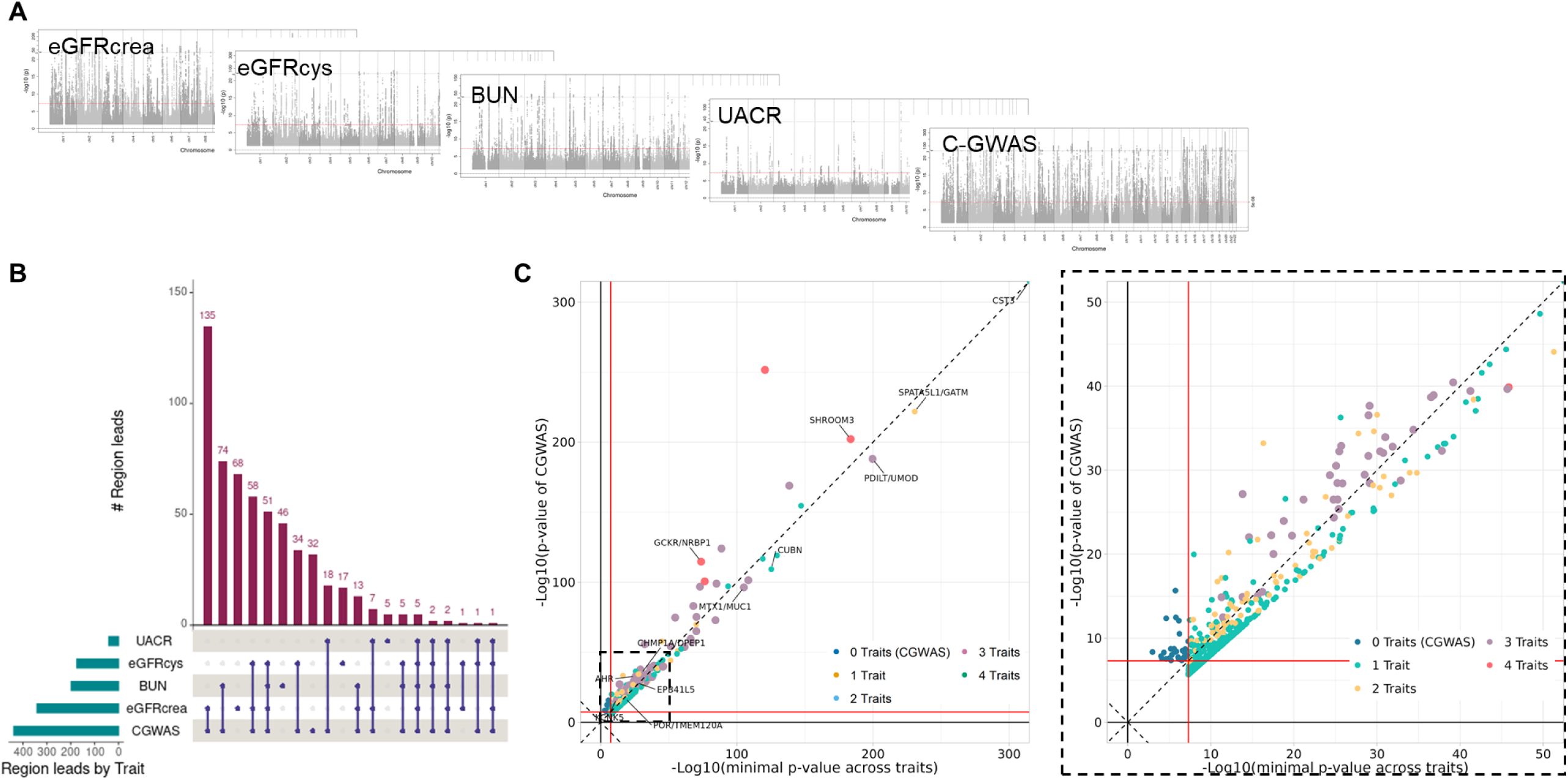
Multi-trait GWAS improves detectability of signals for kidney function traits. **A**: Meta-analysis summary statistics from CKDGen and UKB for eGFRcrea (N=1,004,040), eGFRcys (N=460,826), BUN (N=679,532), UACR (N=547,361) were used as input for C-GWAS (detailed results shown in **Supplementary** Figure 2). **B**: The upset plot is based on the 575 region lead variants identified with genome-wide significance by C-GWAS or any of the four single trait GWAS based on a distance criterion (d>500kb between regions) and depicts the traits for which the specific lead variant was genome-wide significant (P<5×10^-8^). **C**: Shown is a comparison of C-GWAS association P-Value versus the minimum P-values across eGFRcrea, eGFRcys, BUN and UACR. Shown is also a zoomed-in version in the second. The dots are coloured by number of associated traits. Dark blue dots depict region lead variants that were only significantly associated in C-GWAS. The red lines indicate the threshold of genome-wide significance (P<5×10^-8^). P-values were log-transformed for better visibility. Genes known for their involvement with kidney traits and genes discussed in this paper are highlighted.

To identify independent signals at the 575 regions across traits with different sample sizes, we developed a multi-trait conditional analysis approach that combined approximate conditional analyses using GCTA^19^ across multiple trait summary statistics, which was unbiased towards sample size (here referred to as “MT-GCTA”, **Supplementary Figure 4, Supplementary Note 1**, **Methods**): a selection based on P-values would enrich for traits with large sample size; thus, we selected variants that explained most of the trait variance across traits as “index” variant of the signal. Our MT-GCTA approach yielded 812 independent signals (fully conditioned on other signal index variants in the association region: P_c_<5×10^-8^ for at least one trait or for C-GWAS, **Supplementary Table 3**).

In summary, multi-trait GWAS identified 812 independent signals and corresponding signal index variants that were associated with one to four of the kidney function traits at genome-wide significance.

### Multi-trait fine-mapping improves identification of likely causal variants and genes

Next, we assessed whether considering multiple traits helped identify likely causal variants. Bayesian fine-mapping yields a posterior inclusion probability (PIP) for each variant, which can be interpreted as the probability that the variant is causal for the association signal. We conducted multi-trait fine-mapping using flashfm^13^, a method that takes single-trait fine-mapping results as input and refines these by incorporating information from additional traits (**Methods**). We compared the flashfm-refined 95% credible sets with sets from single-trait fine-mapping^20^. While multi-trait fine-mapping is expected to improve fine-mapping for signals that are associated with multiple traits, no improvement is expected for signals exclusively associated with a single trait. We thus applied flashfm to the 325 of the 812 signals that were associated with at least two of the four kidney traits (using P_c_<1×10^-6^ as recommended by the flashfm developers). Across the 325 signals and up to four traits, a total of 774 trait-specific credible sets were considered as input and refined by flashfm (**Supplementary Table 4**). We observed a substantial improvement in fine-mapping by the multi-trait method: credible set sizes were reduced compared to single-trait fine-mapping (median of 6 versus 16 variants); the number of credible sets containing only one variant was substantially increased (161 versus 78, **Figure 2A**). This means that flashfm identified 161 variants that are causal for the association signal with more than 95% probability, while single trait fine mapping only identified 78 of these. The number of credible sets containing a variant with high-PIP (PIP>0.5) increased from 226 for single-trait fine-mapping to 339 for multi-trait fine-mapping. Therefore, 125 high-PIP variants were exclusively identified for the respective trait by the multi-trait method and missed by single-trait fine-mapping (**Figure 2B**).

**Figure 2:**
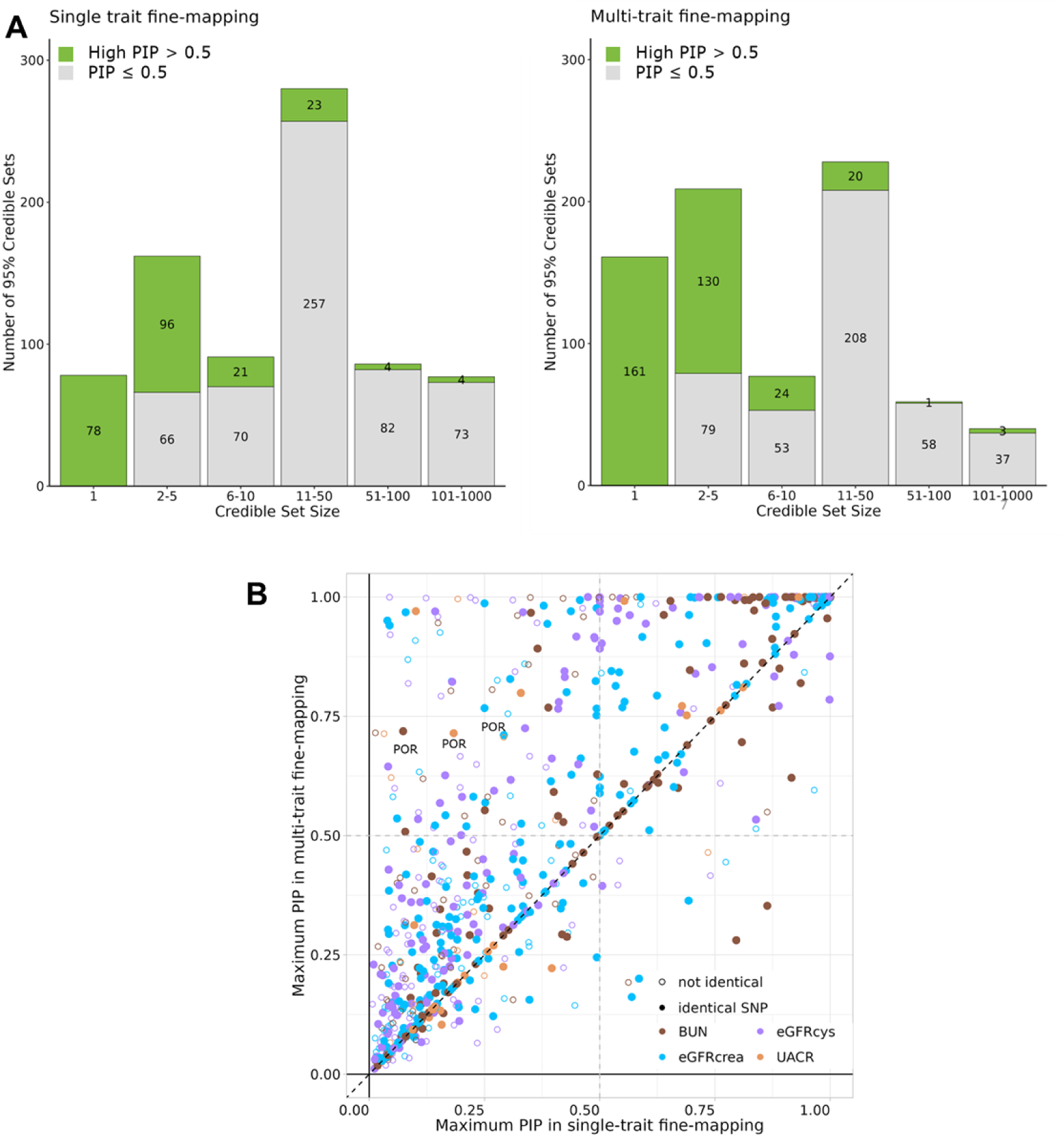
Multi-trait fine-mapping improves identification of likely causal variants. The 812 genome-wide significantly associated (P_c_<5×10^-8^ for any of the four traits or P<5×10^-8^ for C-GWAS) signals were used as input for fine-mapping. A total of 325 of 812 multi-trait signals reached the recommended threshold of P_c_<1×10^-6^ and underwent multi-trait fine-mapping with flashfm^13^. For each of the 325 signals, between two and four credible sets are available depending on the number of traits associated. This resulted in 774 trait-wise 95% credible sets shown below. **A**: Shown are the results separated by set size for single trait fine-mapping (for signals that were also analysed in multi-trait fine-mapping) on the left, and the results of multi-trait fine-mapping and on the right. Coloured in green are variants that have a high-PIP (PIP>0.5) in their respective set. **B**: Shown is the maximum PIP for single and multi-trait fine-mapping for each of the 774 credible sets. Points are coloured by the trait of the credible set. If the variant with the highest PIP is the same in both single- and multi-trait fine-mapping, it is denoted as a filled dot.

To identify likely causal genes around these 125 high-PIP variants, we annotated the variants’ functional consequence^21^ (missense, using dbSNP version 151) or effect on gene expression in kidney tissues^22^ (eQTL in kidney, using NephQTL2, **Methods**). We observed 26 variants that were functionally or regulatory relevant for a total of 39 likely causal genes (**Table 1**, **Supplementary Table 5**). These included eight genes that were only mapped by multi-trait fine mapping (and not by any of our single-trait fine-mapping analyses): *SLC34A1* (for eGFRcrea, eGFRcys and BUN), *MDH2*, *TMEM120A* and *POR* (for eGFRcrea, BUN and UACR), *EEF1AKMT2* (for eGFRcys and BUN), *OAF* (for BUN and UACR), *RERG* (for eGFRcrea, eGFRcys and BUN) and *APH1B* (for eGFRcrea and eGFRcys). Of these, *OAF*, *SLC34A1* and *EEF1AKMT2* have been highlighted previously^9,11,23,24^.

**Table 1.** Likely causal genes exclusively identified by multi-trait fine-mapping. Multi-trait fine-mapping by flashfm yielded 125 trait-specific credible sets with high-PIP (PIP>0.5) variants that were missed by single trait fine-mapping of eGFRcrea, eGFRcys, BUN and UACR (i.e., contained no high-PIP variant in single-trait fine-mapping). Shown here are most likely causal genes that were mapped via these high-PIP variants. Using annotation data describing functional (missense, from dbSNP^21^ version 151) or regulatory relevance (eQTL in kidney tissues from NephQTL2^22^), we found 26 variants and 39 mapped genes. eQTL data is separated for glomerular and tubular-interstitial tissues. The last two columns indicate for which trait the variant was identified as likely causal in single-trait or multi-trait fine-mapping.

| Region ID | Signal ID | Rsid | Chr | Pos | Missense | Glomerular eQTL | Tubular-interstitial eQTL | Single-trait & Multi-trait fine-mapping | Multi-trait fine-mapping only |
| --- | --- | --- | --- | --- | --- | --- | --- | --- | --- |
| *Genes mapped exclusively by flashfm for any trait:* |  |  |  |  |  |  |  |  |  |
| 8 | 1 | rs55785724 | 5 | 176,817,583 |  |  | SLC34A1 | - | eGFRcrea, eGFRcys, BUN |
| 144 | 1 | rs1057868 | 7 | 75,615,006 | POR |  | MDH2, TMEM120A | - | eGFRcrea, BUN, UACR |
| 44 | 1 | rs12247543 | 10 | 126,419,743 |  |  | EEF1AKMT2 | - | eGFRcys, BUN |
| 214 | 1 | rs508205 | 11 | 120,057,343 |  |  | OAF | - | BUN, UACR |
| 45 | 1 | rs10846156 | 12 | 15,321,921 |  |  | RERG | - | eGFRcrea, eGFRcys, BUN |
| 101 | 1 | rs745338 | 15 | 63,598,430 |  |  | APH1B | - | eGFRcrea, eGFRcys |
| *Genes mapped exclusively by flashfm for UACR:* |  |  |  |  |  |  |  |  |  |
| 13 | 1 | rs760077 | 1 | 155,178,782 | MTX1 | MUC1 | MUC1, THBS3 | eGFRcys, BUN | UACR |
| 2 | 1 | rs4859682 | 4 | 77,410,318 |  |  | SHROOM3 | eGFRcrea | eGFRcys, BUN, UACR |
| 38 | 1 | rs4794814 | 17 | 37,696,852 |  |  | FBXL20, PGAP3 | eGFRcrea | BUN, UACR |
| 428 | 1 | rs3814995 | 19 | 36,342,212 | NPHS1 | APLP1 |  | eGFRcrea | UACR |
| *Genes mapped exclusively by flashfm for eGFRcrea or eGFRcys or BUN:* |  |  |  |  |  |  |  |  |  |
| 34 | 1 | rs198325 | 1 | 150,937,405 |  |  | RPRD2 | eGFRcys, BUN | eGFRcrea |
| 13 | 3 | rs1052053 | 1 | 156,202,173 | PMF1-BGLAP, PMF1 |  | PMF1 | eGFRcys | BUN, eGFRcrea |
| 87 | 1 | rs78444298 | 1 | 184,672,098 | EDEM3 |  |  | eGFRcys | eGFRcrea, BUN |
| 82 | 1 | rs11123169 | 2 | 113,967,075 |  | PAX8-AS1 |  | eGFRcrea, BUN | eGFRcys |
| 175 | 4 | rs6555317 | 5 | 498,235 |  | EXOC3 | EXOC3 | eGFRcrea | eGFRcys |
| 175 | 1 | rs13159523 | 5 | 676,962 |  |  | TPPP | eGFRcrea | eGFRcys |
| 17 | 2 | rs7742789 | 6 | 43,345,803 |  |  | CRIP3, TTBK1 | eGFRcys | eGFRcrea, BUN |
| 39 | 1 | rs3757387 | 7 | 128,576,086 |  | IRF5 | TNPO3, IRF5, TSPAN33 | eGFRcrea | eGFRcys |
| 10 | 1 | rs3184504 | 12 | 111,884,608 | SH2B3 |  |  | eGFRcrea | eGFRcys |
| 31 | 1 | rs9529913 | 13 | 72,345,089 |  |  | DACH1 | BUN | eGFRcrea |
| 263 | 1 | rs56261560 | 14 | 99,750,386 |  | DEGS2 |  | eGFRcys | eGFRcrea |
| 106 | 1 | rs11856921 | 15 | 39,278,107 |  |  | THBS1, ENSG00000259345 | eGFRcrea | eGFRcrea |
| 12 | 2 | rs2472297 | 15 | 75,027,880 |  |  | RPP25, MPI | eGFRcrea, BUN, UACR | eGFRcys |
| 212 | 1 | rs34914463 | 17 | 7,366,619 | ZBTB4 |  | CD68 | BUN | eGFRcys |
| 25 | 1 | rs8101667 | 19 | 33,402,419 |  | SLC7A9 | SLC7A9 | eGFRcrea | eGFRcys |
| 81 | 1 | rs429358 | 19 | 45,411,941 | APOE |  |  | eGFRcrea, eGFRcys | BUN |

In summary, our multi-trait fine-mapping substantially improved fine-mapping precision and allowed to resolve kidney trait association signals to their likely causal genes that were missed by single trait fine-mapping.

### Classification yields largely distinct genetics between filtration function and albuminuria with a small overlap

To classify signals as associated with either filtration function or levels of albuminuria or both, we utilized the UKB data on eGFRcrea, eGFRcys, UACR, urinary albumin (Ualb), and urinary creatinine (Ucrea, N up to 436,765, **Methods**). Focussing on UKB allowed for a fair comparison of genetic effect sizes and their precision due to comparable sample size and largely the same individuals across traits. For this, we re-estimated fully conditional genetic effects on each trait for the 812 independent signal index variants that were genome-wide significant in the meta-analyses. Of these 812 variants, 740 variants showed significant association in UKB (fully conditioned P_c_<0.05/812 in UKB for at least one trait, **Supplementary Table 6**). These were considered for further classification.

First, we ascertained the genetic variant relevance for filtration function (“real eGFR”) opposed to mere biomarker metabolism by joint effects on eGFRcrea and eGFRcys and relevance for “real UACR” opposed to mere Ucrea associations by joint effects on UACR and Ualb (**Methods**). Among the 740 variants, 436 were associated with eGFRcrea or eGFRcys (any P_c_<0.05/740) comprised of (i) 308 variants classified as “real eGFR” (both eGFRcrea and eGFRcys P_c_<0.05/740, directional consistent), (ii) 90 variants as creatinine metabolism (“eGFRcrea-only”, eGFRcrea P_c_<0.05/740 and eGFRcys P_c_>0.05), and (iii) 38 variants as cystatin C metabolism (“eGFRcys-only”, eGFRcys P_c_<0.05/740 and eGFRcrea P_c_>0.05; **Figure 3A, Supplementary Table 6**). Three variants were associated with both eGFRcrea and eGFRcys but with opposite direction (including one near *CPS1*). Most of the 308 “real eGFR” variants demonstrated significant association with BUN in the expected direction (94.3%, i.e., opposite direction for BUN compared to eGFR; **Supplementary Figure 5**).

**Figure 3.**
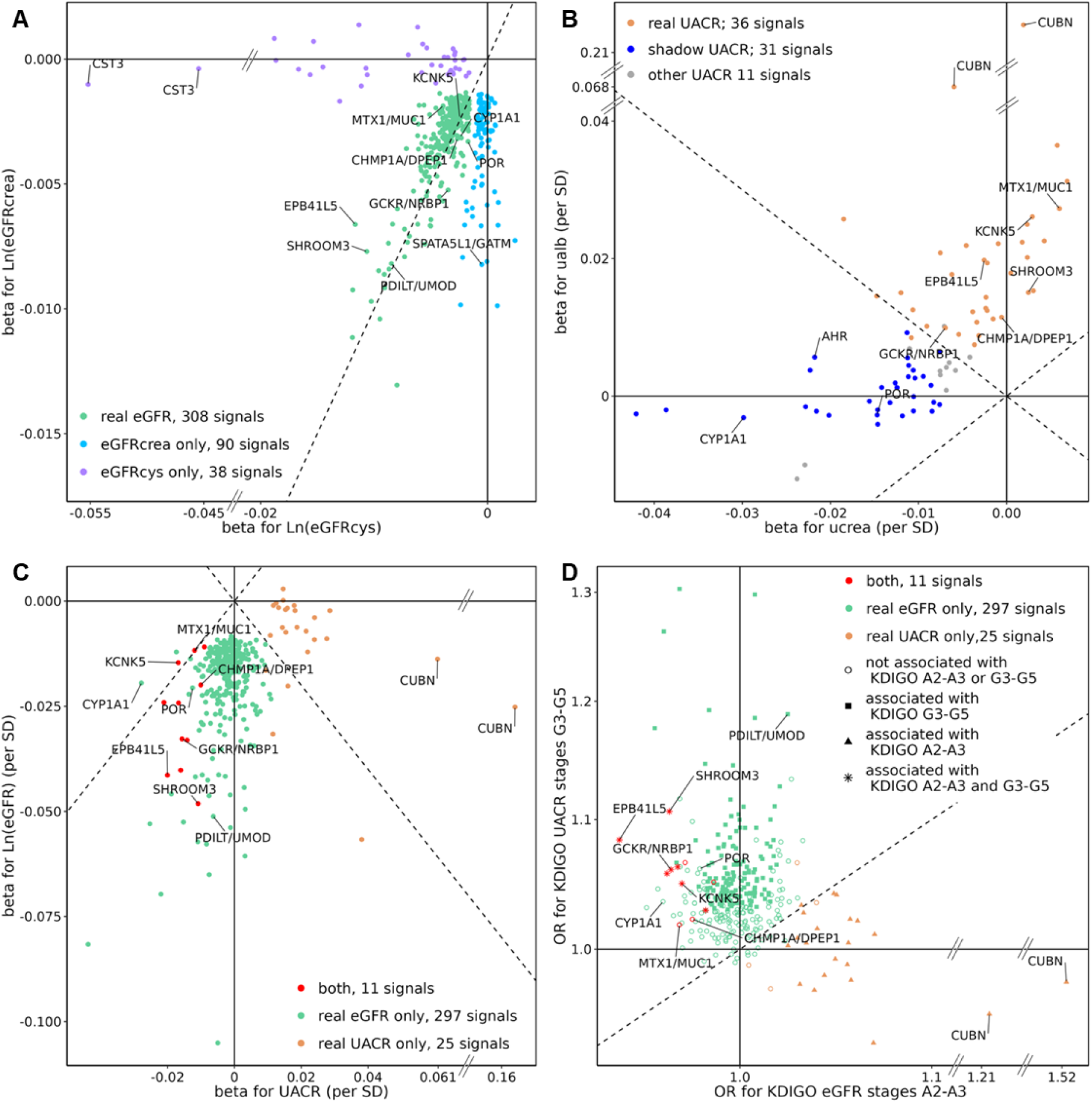
Classification of kidney trait signals yields distinct genetics of “real eGFR” versus “real UACR” with little overlap. In UKB, 740 signals were associated with any of the four traits (P_c_<0.05/812, eGFRcrea N=436,581, eGFRcys N=436,765, BUN N=435,677, or UACR N=444,861). Effect alleles were turned to match eGFRcrea decreasing alleles, except for B and C “real UACR only”, which were turned to match UACR increasing alleles **A**: Among the 740 variants, 436 were associated with eGFRcrea or eGFRcys (P_c_<0.05/740). We classified these regarding “real eGFR” (eGFRcrea and eGFRcys P_c_<0.05/740, consistent direction, green), “eGFRcrea-only“ (eGFRcrea P_c_<0.05/740 and eGFRcys P_c_>0.05, blue) and “eGFRcys-only“ (eGFRcys P_c_<0.05/740 and eGFRcrea P_c_>0.05, purple). Effect sizes shown are on log-transformed eGFRcrea and eGFRcys. **B**: Among the 740 variants, 78 were associated with UACR (P_c_<0.05/740). We separated these into variants associated with increased Ualb (N=444,861, “real UACR”, P_c_<0.05/78, orange), variants associated with increased Ucrea (N=444,861, “shadow UACR”, P_c_<0.05/78, blue) and the rest (grey, “other UACR”). Effect sizes are on inverse-normal transformed Ualb and Ucrea. **C**: Shown are standardized effect sizes on eGFR (using eGFRcrea or eGFRcys, based on stronger association) versus UACR for 333 variants associated with “real eGFR” or “real UACR” (SD_eGFRcrea_=0.16, SD_eGFRcys_=0.2, SD_UACR_=1) and their classification (“real eGFR only”: green; “both”: red, “real UACR only”: orange). **D**: For the 333 variants, shown are odds ratios with clinical outcomes according to KDIGO stages G3-G5 or A2-A3 (eGFR<60ml/min/1.73² or UACR≥30mg/g, respectively, P_c_<0.05).

Second, among the 740 variants, 78 were associated with UACR (P_C_<0.05/740), which included 36 variants ascertained for directional consistent association with Ualb (Ualb P_C_<0.05/78; “real UACR”) and 31 not ascertained for Ualb but for Ucrea (Ucrea P_C_<0.05/78 and Ualb P_C_>0.05/78, “shadow UACR”, **Figure 3B, Supplementary Table 6**). Thus, 36 variants showed evidence for association with albumin leakage into urine rather than a shadow association with UACR merely by association with the denominator Ucrea. Among the 31 “shadow UACR” effects were 25 variants that were not associated with real eGFR. Thus, these 25 variants were identified only by association with Ucrea (“shadow UACR only”).

Finally, we were interested in the overlap of “real eGFR” genetics with “real UACR” genetics, as done previously by Ellis and colleagues^12^, but now with a much larger sample size (N up to 444,861 compared to 31,580 from Ellis et al.). Among the 333 variants identified for “real eGFR” or “real UACR”, we found that (i) 297 were ”real eGFR only”, (ii) 25 were “real UACR only”, and (iii) 11 were “both” (**Figure 3C, Supplementary Table 6**). For the 11 variants, all the eGFR-lowering alleles were associated with decreased UACR (**Table 2**). The 11 variants included one signal near *SHROOM3*, for which this pattern was reported previously^12^, and ten novel such signals. Gene mapping based on high-PIP variants at these signals yielded 4 genes mapped by missense variants (*EPB41L5, GCKR, MTX1 and THBS3*) and 8 mapped by kidney tissue eQTLs (*CHMP1A, DPEP1, GTF3C2-AS2, LINC02166, MUC1, NRBP1, SHROOM3,*

**Table 2.** Eleven signals associated with “real eGFR” and with “real UACR”. Shown are 11 signal index variants that were associated with “real eGFR” (eGFRcrea N=436,581; eGFRcys N=436,765) and with “real UACR” (UACR N=444,861; Ualb N=444,861) in UKB. Effect estimates are fully conditioned and standardized using SD_eGFRcea_=0.16, SD_eGFRcys_=0.2, SD_UACR_=1, SD_ualb_=1, SD_ucrea_=1 (Ucrea from UKB, N=444,861). Bonferroni-corrected significant associations are marked in bold (P_c_<0.05/740 for eGFRcrea, eGFRcys and UACR, P_c_<0.05/78 for Ualb). High-PIP variant RSIDs are marked in bold. Additionally, missense variants (dbSNP version 151, Phan et al.,2025) and regulatory relevant variants are indicated by the influenced gene, separated by kidney tissue type^22^. The effect direction of eQTL estimates is indicated by (-) and (+).

| RSID | Nearest Gene | EA | eaf | eGFR<sub>crea</sub> |  | eGFR<sub>cys</sub> |  | UACR |  | Ualb |  | Ucrea |  | Missense | Gene mapping |  |
| --- | --- | --- | --- | --- | --- | --- | --- | --- | --- | --- | --- | --- | --- | --- | --- | --- |
|  |  |  |  | beta | P | beta | P | beta | P | beta | P | beta | P |  | Glomerular eQTL | Tubular-interstitial eQTL |
| rs1611774 | TRIM46 | t | 0.1 | -0.02 | **1.3E-09** | -0.02 | **2.1E-08** | -0.02 | **6.2E-07** | -0.015 | **8.0E-05** | 0.012 | 0.0044 | THBS3 | - | - |
| **rs760077** | MTX1 | a | 0.4 | -0.01 | **4.1E-08** | -0.02 | **9.1E-22** | -0.01 | **1.6E-07** | -0.027 | **2.5E-40** | -0.006 | 0.0085 | MTX1 | MUC1(-) | MUC1(-), THBS3(-) |
| rs4442348 | LOC100422212 | g | 0.5 | -0.01 | **4.6E-08** | -0.01 | **5.7E-13** | -0.01 | **2.4E-05** | -0.008 | **9.5E-05** | 0.0037 | 0.084 |  | - | - |
| **rs1260326** | GCKR | c | 0.6 | -0.03 | **5.1E-55** | -0.02 | **5.2E-18** | -0.02 | **2.1E-12** | -0.01 | **8.4E-07** | 0.007 | 0.0018 | GCKR | - | GTF3C2-AS2(-), NRBP1(-) |
| **rs28930677** | EPB41L5 | t | 0.1 | -0.04 | **1.6E-18** | -0.06 | **3.5E-38** | -0.02 | **6.7E-05** | -0.02 | **1.2E-05** | 0.0026 | 0.61 | EPB41L5 | - | - |
| rs28394165 | SHROOM3 | c | 0.5 | -0.05 | **1.1E-114** | -0.05 | **4.2E-152** | -0.01 | **1.5E-06** | -0.015 | **1.2E-13** | -0.003 | 0.27 |  | - | SHROOM3<sup>#</sup> (+) |
| **rs80138475** | ARL15 | c | 0.9 | -0.04 | **5.7E-37** | -0.03 | **1.9E-16** | -0.02 | **1.9E-06** | -0.019 | **2.0E-10** | 0.0022 | 0.51 |  | - | - |
| rs62398607 | KCNK5 | t | 0.8 | -0.02 | **2.3E-09** | -0.01 | **1.7E-07** | -0.02 | **1.2E-10** | -0.026 | **1.6E-28** | -0.003 | 0.26 |  | - | - |
| rs881858 | LOC100132354 | a | 0.7 | -0.03 | **3.9E-53** | -0.03 | **1.6E-56** | -0.01 | **6.4E-10** | -0.014 | **3.7E-12** | 0.0023 | 0.31 |  | - | - |
| **rs28601761** | TRIB1 | g | 0.4 | -0.02 | **1.2E-32** | -0.01 | **2.4E-08** | -0.02 | **1.3E-14** | -0.009 | **1.5E-05** | 0.011 | **5.9E-07** |  | - | - |
| rs164747 | CHMP1A | g | 0.4 | -0.02 | **6.4E-23** | -0.01 | **6.3E-12** | -0.01 | **3.3E-06** | -0.011 | **3.8E-09** | 0.0006 | 0.79 |  | DPEP1* (+) | DPEP1*(+), CHMP1A* (-), LINC02166* (+) |

<sup>#</sup> SHROOM3 was mapped by high-PIP variant rs4859682, which is a perfect proxy to the index variant rs28394165 ( $r^2=1.0$ in EUR from 1000 Genomes); \* eQTLs for signal index variant rs164747 $PIP_{eGFR_{crea}}=0.43$ , $PIP_{eGFR_{cys}}=0.06$ , $PIP_{UACR}=0.02$ , only single-trait fine-mapping was performed.

*THBS3*, **Table 2**). These 11 signals benefited particularly from multi-trait fine-mapping, since they required association with eGFRcrea, eGFRcys and UACR (median credible set size reduced from 6 to 2; number of high-PIP variants increased from 5 to 8). Specifically, rs4859682 a tubular-interstitial eQTL for *SHROOM3*, demonstrated high-PIP for eGFRcrea in single- and multi-trait fine-mapping (PIP>95%), but for eGFRcys and UACR only in flashfm (PIP=0.96 and 0.97, respectively; 0.33 and 0.10 with single-trait fine-mapping). Multi-trait fine-mapping also sharpened signals in “real eGFR only”, which required eGFRcrea and eGFRcys associations (median set size: 15 to 5; number of high-PIP variants: 98 to 106), but did not improve “real UACR-only”.

Since quantitative eGFR and UACR in general population reflects mostly physiological levels, we were interested whether the 11 variant association with both traits also holds for KDIGO stages G3-G5 or stages A2-A3 (eGFR<60 ml/min/1.73m² or UACR≥30 mg/g, respectively; logistic regression, **Methods**): all 11 variants showed odds ratios directionally consistent with the quantitative trait associations, including seven that were nominally significant for both binary outcomes (P_c_<0.05, near *SHROOM3*, *GCKR, EPB41L5, ARL15, KCNK5, TRIB1 and LOC100422212*; **Figure 3D**).

In summary, this illustrates the gain from exploiting multi-trait associations to help identify likely causal variants and genes for “both” and “real eGFR only” signals. We found largely distinct genetics of “real eGFR” and “real UACR” with a small overlap: 297 signals for “real eGFR only”, 25 for “real UACR only”, and 11 for “both”. While one signal, near *SHROOM3*, has been previously described for the eGFR&UACR overlap^12^, we here found 11 such signals including *SHROOM3* and all 11 eGFR-lowering alleles were associated with decreased UACR.

### Kidney cell-type specific expression patterns are undifferentiated by class

Next, we were interested whether mapped genes by class pinpointed distinct biology regarding the involved kidney tissue and cell-type. When querying the 123, 8, and 11 high-PIP variants (PIP>0.5 from multi-trait or single-trait fine-mapping) among “real eGFR only”, “both”, or “real UACR only” signals, respectively, for being protein-altering or eQTL in glomerular or tubular-interstitial tissues, we mapped 56, 11, and 10 likely causal genes (**Figure 4, Supplementary Table 7**). Of the 3 genes mapped at “shadow UACR only” signals, none were mapped by an eQTL in kidney tissue.

**Figure 4:**
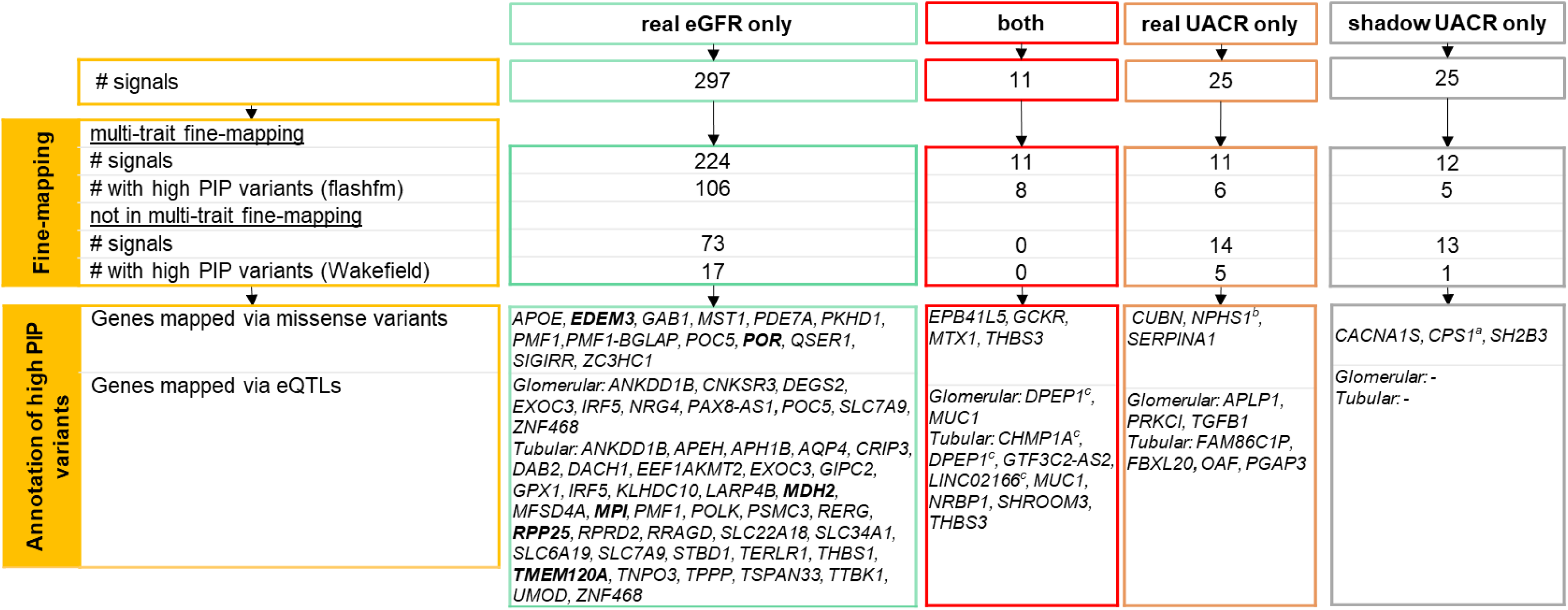
Class-specific fine-mapping and gene mapping yield likely causal genes. In this summary, we show, for each class, the number of signals, a summary of fine-mapping results and mapped genes. For fine-mapping, multi-trait fine-mapping by flashfm was applied when at least two traits displayed association at P_c_<10^-6^. Other signals were fine-mapped based on the Wakefield method. Genes were mapped to high-PIP variants using functional annotations by dbSNP version 151^21^ and by eQTLs^22^ in relevant tissues. “Real eGFR” genes that also demonstrated “shadow UACR” effects are marked in bold. For example, *POR* was classified as “real eGFR” and also as “shadow UACR”.

At signals involving “real eGFR” (“real eGFR only”, “both”), most genes were mapped by high-PIP eQTL in tubular-interstitial tissue, while both tissue types appeared similarly for “real UACR only” (**Figure 5A**). The most recognized representatives of each of the classes, “real eGFR”, “both”, or “real UACR” were *UMOD*, *SHROOM3*, and *CUBN*, respectively.

**Figure 5:**
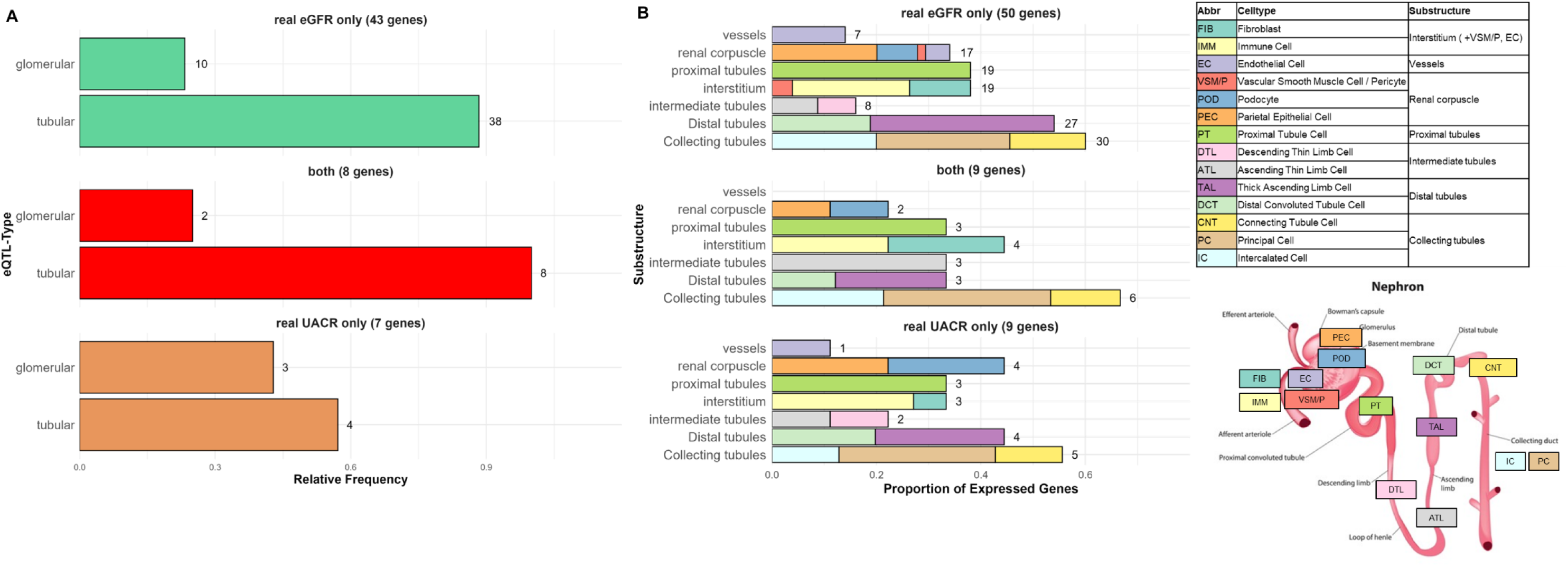
Gene mapping yielded shared kidney cell-type expression patterns. Genes of the classes “both”, “real eGFR only” and “real UACR only” (Figure 4) were queried for differential kidney cell-type specific expression from the KPMP kidney tissue atlas^25^. **A**: Shown are the relative number of eQTL-mapped genes in glomerular or tubular-interstitial tissue among all eQTL-mapped genes. **B**: Shown is the distribution of cell types in the substructures sorted by classes. Shown is also a nephron marked with different cell types matching the substructures and the descriptive table (nephron picture from https://www.shutterstock.com/image-vector/biological-structure-nephron-diagram-600nw-2295097935.jpg).

When querying mapped genes regarding cell-type specific differential expression using the kidney cell atlas^25^ (**Methods**), we found differential expression in cell-types of all sections of the nephron with little distinction across classes (**Figure 5B**). The five novel mapped genes (*MDH2*, *TMEM120A*, *POR*, *RERG*, and *APH1B*) showed absolute and differential expression in a variety of cell types^26^ (**Supplementary Figure 6+7A**). Differential and absolute expression was mostly pronounced in tubular cells across classes, but also in podocytes (*NPHS1, APLP1, PRKCI*, *SHROOM3*, *EPB41L5*, *THBS1, RERG, DAB2, PAX8-AS1, DACH1*). The strongest differential expression in podocytes was found for *NPHS1* (**Supplementary Figure 6+7B**) well-known for Mendelian disease with damaged podocyte structure^27^. Our missense variant mapping to *NPHS1* (rs3814995) was a “real UACR only” signal but was also directionally concordantly associated with eGFRcrea and just missed as a “both” signal due to a weak eGFRcys association (P=1.5×10^-14^, 0.006, and 4.2×10^-5^ for eGFRcrea, eGFRcys, and UACR, respectively).

Overall, the tissue- and cell-type-specific expression pattern supported the mapped genes’ role in all sections of the nephron un-differentiated by class.

### Genetics of UACR without albumin leakage into urine is linked to fluid intake genetics

To better understand the etiology of shadow associations on UACR by mere association with Ucrea without evidence for albumin leakage into urine (no association with Ualb), we evaluated a potential role of fluid intake. Our GWAS in UKB on fluid intake volume (from water, coffee or tea, N=357,00, N=358,093 N=373,481, **Methods**) identified 88 independent signals (**Supplementary Figure 8A**). Strongest associations were around *CYP1A2/A1, POR* and *AHR* known for their association with caffeine intake^28–30^. However, their coffee/tea-intake increasing alleles were also associated with increased total fluid intake (**Supplementary Table 8**). This is in line with increased caffeine preference leading to increased total fluid intake. Genes in the association regions were enriched for differential expression in brain (FUMA, **Supplementary Figure 8B**). Fluid-intake increasing alleles decreased Ucrea, which is in line with decreased urine concentration by increased fluid intake (**Supplementary Figure 8C**).

Among the 31 “shadow” UACR variants (driven by Ucrea associations), 9 were associated with fluid intake (P<0.05/31), where fluid-intake increasing alleles increased UACR (**Figure 6A**, **Supplementary Table 9**). Mediation analyses demonstrated that Ucrea effects near *CYP1A1*, *POR* and *AHR* were mediated by fluid intake to 40-50% (**Figure 6B**, **Methods**). Given a considerable measurement error of fluid intake, this could be indicative of an even substantially larger mediation. Noteworthy, 4 of the 9 variants had effects on “real eGFR”, and eGFR-effects were not mediated by fluid intake suggesting independent effects on caffeine preference and filtration rate (**Figure 6C, Supplementary Table 9**). For example, the lead variant of the signal around *POR*, a high-PIP variant, is missense for *POR* and is an eQTL for *MDH2*/*TMEM120A* in kidney tissue, which indicates behavioural effects of the variant on caffeine preference independent of physiological effects on kidney-related mechanism affecting filtration rate^31,32^ (**Supplementary Note 2**).

**Figure 6:**
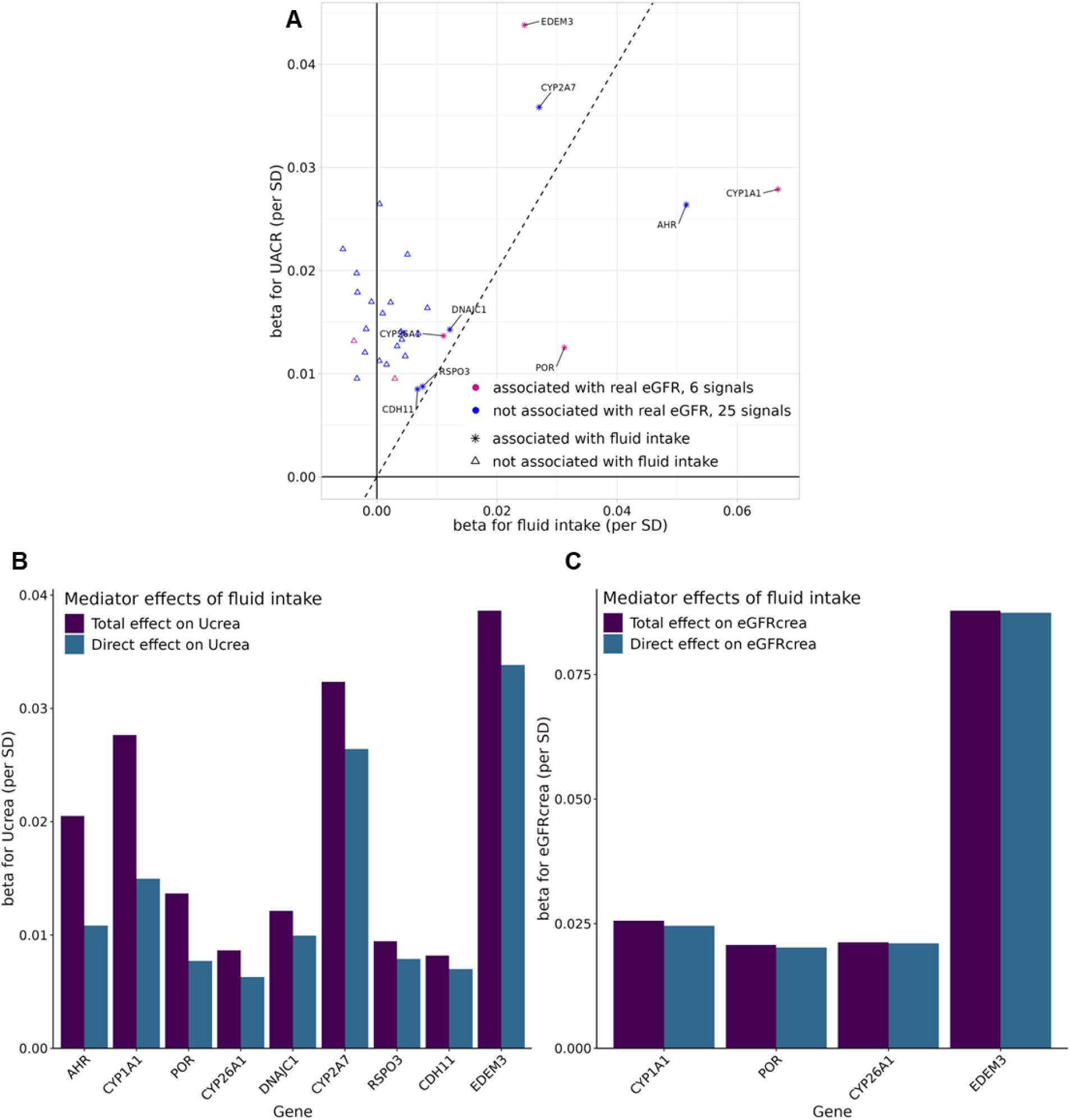
“Shadow UACR” variants are linked to fluid intake and to “real eGFR”. For the 31 “shadow UACR” variants, we assessed their association on fluid intake and on eGFR and investigated whether fluid intake mediated their effects on UACR and eGFR**. A**: For the 31 “shadow UACR” variants, fully conditioned and standardized effect estimates (SD_UACR_=1, SD_fluid intake_ =0.77) effect estimates for UACR and fluid intake are shown. Symbols depict the association status (P_C_<0.05/31) of the variant with fluid intake. **B**: For the 9 signals associated with fluid intake, mediator analyses were conducted to investigate whether their effects on Ucrea are mediated by fluid intake. Shown are total and direct (mediator-adjusted for fluid intake) effects of the index variants on Ucrea. **C**: For 4 of the 9 variants that were further associated with “real eGFR”, mediator analyses were performed to investigate whether their effect on eGFR are mediated by fluid intake. Shown are total and direct (mediator-adjusted for fluid intake) effects of the variants on eGFRcrea.

Thus, this underscores that fluid intake genetics can result in “shadow UACR” effects without evidence for albumin leakage into urine.

### About the interplay of kidney function traits with other kidney-related phenotypes

We aimed to better understand the interplay of kidney function traits with each other and traits related to systemic perfusion pressure, blood sugar control or fluid intake.

First, we evaluated the relationship of eGFR- and UACR-traits with each other. For eGFRcrea and eGFRcys, we observed moderate positive genetic and phenotypic correlations (r_g_=0.60, r_p_=0.58, **Methods**). This indicates a shared etiological basis, but also distinct etiology including biomarker metabolism (**Figure 7A**). For UACR, correlations were high with Ucrea (r_g_=-0.79, r_p_=-0.78), but weak with Ualb (r_g_=0.17, r_p_=0.09, **Figure 7A**). Linking eGFR and UACR, we observed moderate positive genetic correlations for eGFRcrea or eGFRcys with UACR (r_g_=0.36, r_g_=0.23, respectively), despite weak phenotypic correlations (r_p_<0.10, **Figure 7A**). This indicates that eGFR and UACR are largely uncorrelated at the phenotypic level in the rather healthy UKB study population^33^, but positively correlated at the genetic level. This positive correlation derives from genetic variants like our “both” class variants or from genetic variants associated with eGFR and “shadow UACR” effects.

**Figure 7:**
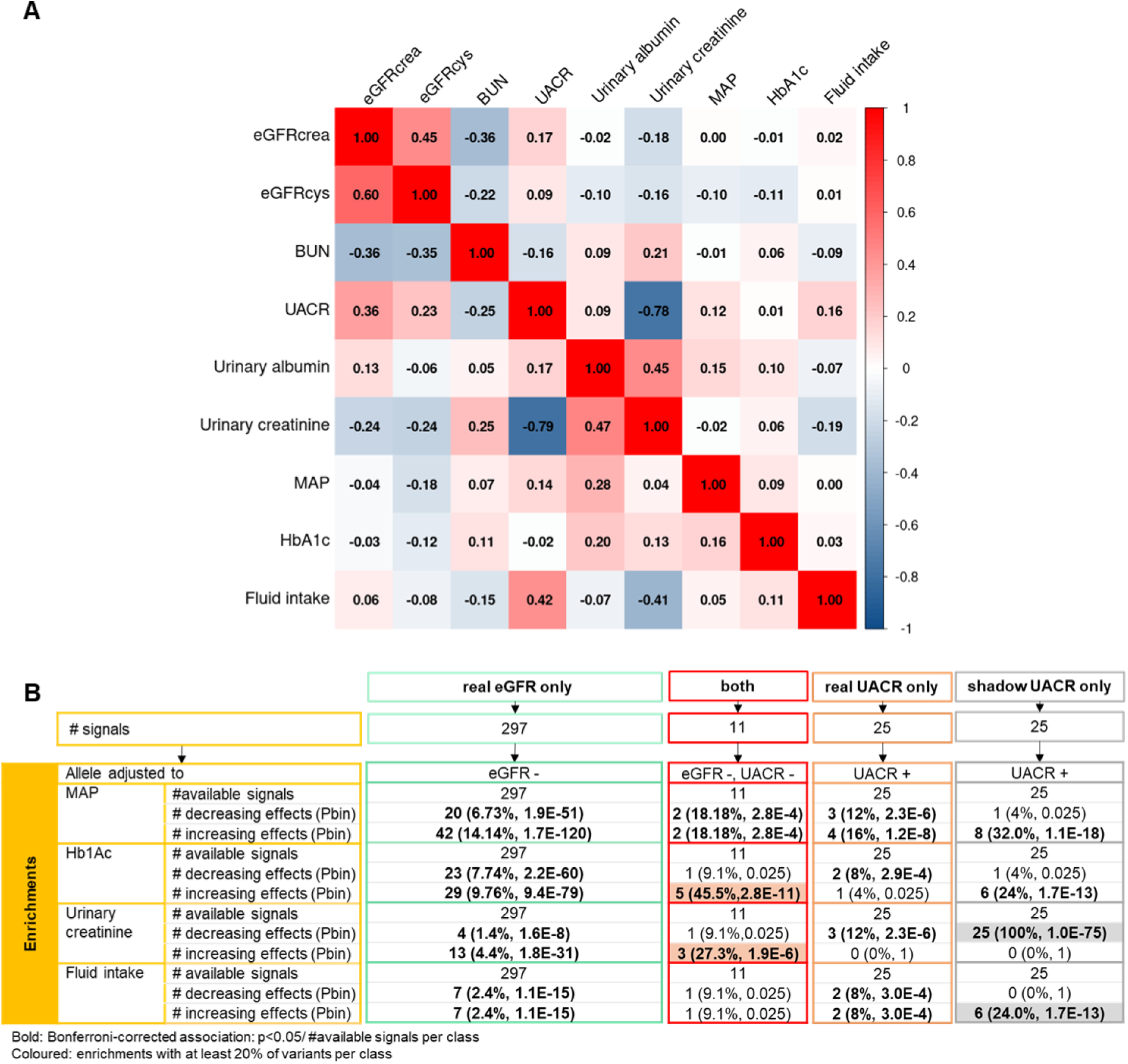
General and class-specific interplay of kidney-related traits. To investigate the interplay of primary and secondary traits using UKB, we conducted correlation analyses (irrespective of class) and enrichment analyses (by class). **A**: Shown are genetic correlations (lower triangle) based on LD score regression and phenotypic correlations (Spearman, upper triangle) for 11 kidney related traits using GWAS summary statistics from UKB (N up to 436K). For phenotypic correlation all traits were adjusted for age and sex to ensure comparability. **B**: One-sided binomial enrichment of significant effects was tested for each class with multiple outcomes. Association was tested as P<0.05/number of signals in class. Binomial enrichment was tested against the same P-value divided by two for one-sided testing. Significant enrichments were marked in bold (P<0.05/number of signals per class) and enrichments containing at least 20% of class variants are coloured.

Second, we explored the relationship of kidney function traits with mean arterial pressure (MAP), a measure of systemic perfusion pressure, glycated haemoglobin (HbA1c), a measure of long-term blood sugar control, and fluid intake. There was little correlation, except a moderate genetic correlation of Ucrea and UACR with fluid intake (r_g_=-0.41, r_g_=0.42, respectively; **Figure 7A**).

Third, we were interested in any pattern of enrichment for MAP, HbA1c or fluid intake associations among the identified variants by class: we found a multitude of enrichments undifferentiated by class, except for a strong enrichment of (i) “both” class variants for HbA1c association (45% of the 11 variants with P<0.05/11) and (ii) “shadow UACR only” variants for Ucrea or fluid intake association (100% or 24%, respectively; **Figure 7B, Supplementary Table 10, Methods**).

Together, the genetic correlation between eGFR and UACR was in line with the existence of “both” class signals and it was the “both” class that was enriched for variant association with HbA1c. However, any observed pattern does not establish causality and may also reflect pleiotropy or complex interplay.

### Statistical and biological characterization reveals distinct mechanisms for “both” class signals

We were particularly interested in helping resolve causality and biology of the 11 “both” class signals and mapped genes.

To statistically disentangle potential causal mechanisms linking eGFR and UACR effects with each other and MAP and HbA1c effects, we conducted a mediation analysis^34^ (**Methods**). Our findings indicate that the effects of most of the 11 genetic variants on eGFR were not or not substantially mediated by their effect on UACR, but their effects on UACR were largely mediated by eGFR (**Figure 8A**, **Supplementary Table 11**). The *SHROOM3* variant exhibited total mediation of the UACR effect by eGFR: there was no direct effect on UACR left after adjustment for eGFR (proportion mediated = 100% for G→eGFR→UACR vs. 2% for G→UACR→eGFR; **Figure 8B**). While some of the 11 variants showed significant associations with MAP or HbA1c (P<0.05/11), none showed mediation by MAP or HbA1c (proportion mediated <10%; **Figure 8B, Supplementary Figure 9**). This suggest that effects were primarily on filtration function with secondary effects on albumin leakage into urine, that are not mediated by MAP or HbA1c.

**Figure 8:**
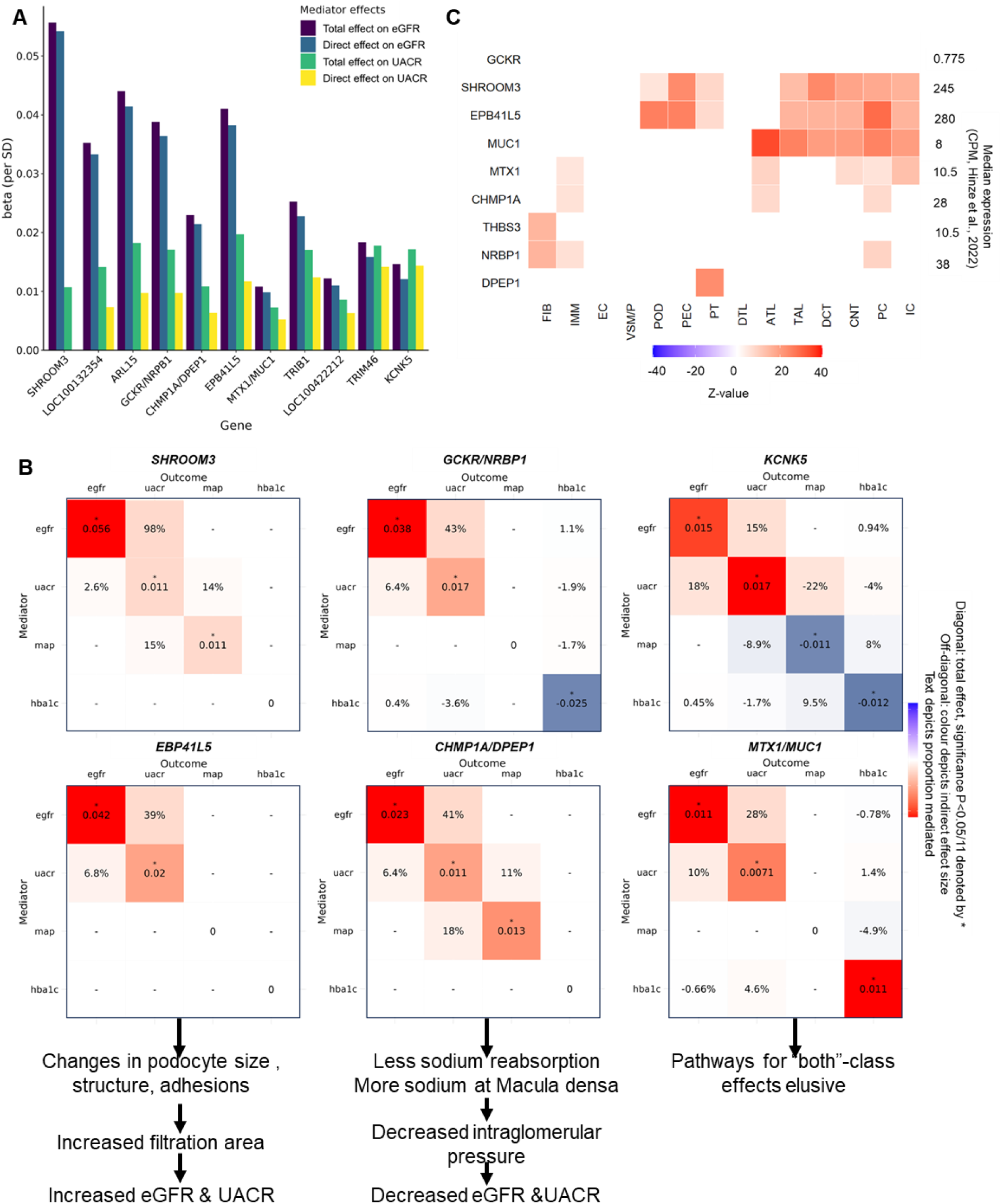
Characterization of both class signals reveals distinct mediation and expression patterns. The 11 “both” class signals were characterized using mediator analyses on signal index variants and queries of differential expression in kidney tissue on mapped genes. **A**: For the 11 ”both” class signals, mediator analyses were conducted to investigate their effects on eGFR and UACR. Shown are total and direct (mediator-adjusted) effects of the index variants on eGFR and UACR. The direct effect on eGFR can be considered as “UACR-adjusted” effect with the indirect effect being the eGFR effect that is mediated through UACR. **B**: The heatmaps show detailed results from mediator analyses on all combinations eGFR, UACR, MAP and HbA1c: On the diagonal is the total genetic effect on the outcome, off-diagonal are the proportions of mediation on the outcome. **C**: For the 11 genes mapped in “both” signals, the heatmap shows cell-type specific expression from KPMP^25^. Any significantly differentially expressed cell-type is coloured. On the right side the median expression in counts per million is depicted. Used were both healthy and injured kidney cells^26^.

When evaluating the 11 genes mapped to the “both” class signals regarding biological mechanisms, we found three types of genes (**Figure 8B**): (i) two genes, *SHROOM3* and *EPB41L5*, showed differential expression in podocytes and distal tubulus (**Figure 8C**), in line with their known effects on podocyte size or adhesion^35,36^, respectively. The *SHROOM3* knock-out is known for podocyte damage in mice^35^ and the eGFR&UACR-increasing allele of the high-PIP variant mapped *SHROOM3* by decreased expression in tubulo-interstitium. This is in line with a hypothesis that the eGFR&UACR-increasing allele decreases *SHROOM3* expression in podocytes, impairs podocyte structure, and increases filtration area. This would yield increased filtration of creatinine, cystatin and albumin (**Supplementary Note 3**). The secondary effect on albumin leakage in urine might be explained by delayed effects when the albumin reuptake starts being exhausted. It might also be explained by secondary causal, less frequent, variants jointly inherited with the high-PIP variant^37^: when the variants make up three sets of haplotypes in *SHROOM3*, where the first two haplotype sets increase filtration area and only the second haplotype set affects tubular reuptake (third haplotype set as reference). This would lead to a mediated UACR-effect when adjusted for eGFR. A similar mechanism might be involved for the *EPB41L5*, where podocyte adhesion^36^ affects filtration area with similar consequences on increased filtration and secondary effects on albumin leakage into urine as for *SHROOM3* (**Supplementary Note 3**). Partial mediation of the UACR-effect when adjusting for eGFR (**Figure 8A/B**) might derive by a haplotype effect as described for *SHROOM3*, where now the haplotypes relevant for tubular reuptake are not a full subset of the haplotypes relevant for the filtration area. (ii) Three further genes, *NRBP1*, and two genes mapped by one signal, *CHMP1A* and *DPEP1*, are linked to sodium-mediated intraglomerular pressure regulation (**Figure 8B**, **Supplementary Note 3**): *NRBP1* affects sodium reabsorption via *WNK* signalling^38^, and the signal affecting *DPEP1/CHMP1A* expression is known for a joint role in iron homeostasis^39^ that could also affect sodium-mediated intraglomerular pressure. The variant that modifies *NRBP1* expression is also missense for *GCKR*, a gene known for diabetes^40^, which is in line with the variant’s effect on HbA1c. However, the observed independence of the variant’s effects on eGFR&UACR from its effect on HbA1c suggests that the diabetes-related effect operates through *GCKR*, while the eGFR&UACR effects derive from *NRBP1* (**Figure 8B**). Together, this prompts a hypothesis that both *NRBP1* and *DPEP1/CHMP1A* signal effects regulate intraglomerular pressure and thus filtration of creatinine, cystatin and albumin with secondary effects on albumin leakage into urine. Our mediator analyses showed that these effects were independent of MAP, which indicates a kidney-intrinsic regulation of glomerular pressure rather than systemic pressure. (iii) For the genes in the other five signals, hypotheses of mechanisms were more elusive. The variant in *MTX1* (missense) also regulates *MUC1* expression and *MUC1* is expressed in many kidney cells (**Figure 8C**). The variant in/near *KCNK5*, known for its role for potassium regulation^41^, showed independent effects on eGFR and UACR as well as independent effects on MAP and HbA1c (**Figure 8B**).

In summary, the 11 concordant eGFR and UACR associations appear to derive, at least in part, from effects on podocyte structure affecting filtration area (*SHROOM3* and *EBP41L5*) or effects on sodium-mediated regulation of intraglomerular pressure (*NRBP1, CHMP1A/DPEP1*). Both increased filtration area or increased intraglomerular pressure would lead to increased filtration of creatinine, cystatin, and albumin. Mediator analyses highlighted that these effects were independent of systemic blood pressure or glucose control. The “both” group genetics thus pinpoints important kidney-intrinsic mechanisms.

## DISCUSSION

Our large-scale multi-trait GWAS, encompassing 4 kidney traits in up to 1 million individuals, and multi-trait fine-mapping enabled the identification of novel likely causal variants and genes for filtration rate and/or albumin leakage into urine. Our results revealed that the genetics of filtration function are largely distinct from the genetics of albuminuria and that there is a small, but physiologically very interesting overlap. We also highlighted genetic variants that showed a shadow effect on UACR by association with Ucrea without any albumin leakage into urine and that a large proportion of these variant effects were mediated by fluid intake.

Our results revealed 11 signals with associations on both eGFR and UACR with a directionally distinguished pattern: the 11 eGFR-increasing alleles were all associated with increased UACR. One such variant, rs17319721, near *SHROOM3*, was previously reported^12^ and highly correlated (r² = 0.90) with our *SHROOM3* lead variant, rs28394165, that is a likely causal variant affecting *SHROOM3* expression in tubular-interstitial tissue. We showed that most of these 11 effects were primary effects on eGFR, that UACR-effects were substantially mediated by these eGFR-effects, and that these effects were independent of MAP or HbA1c. Genes mapped by highly likely causal variants in these 11 signals suggested these effects to derive – at least in part – from increased filtration area from impaired podocyte structure^35,36^ (*SHROOM3* and *EBP41L5*) or from increased glomerular filtration pressure^38,39^ (*NRBP1, CHMP1A/DPEP1*). Both mechanisms would result in increased filtration of creatinine, cystatin and albumin with secondary effects on albumin leakage into urine. These secondary effects on albumin leakage might be delayed effects by albumin re-uptake in tubule or secondary causal variants that independently affect albumin re-uptake^37^ or a combination of both. The concordant effects on eGFR&UACR for *SHROOM3* were hypothesized to be pleiotropy or albuminuria in hyperfiltration states, including diabetes, rare kidney diseases or hypertension^12^. Our mediator analyses rule out that the genetic variants cause elevated systemic perfusion pressure or impaired glucose control with hyperfiltration as consequence, and only few individuals in UK Biobank have manifest rare kidney diseases. The concordant effects might rather be viewed as a physiological effect, in comparably healthy individuals, on increased filtration followed by albumin leakage into urine. Overall, the concordant effects can be considered “pleiotropy” in the sense of mediated pleiotropy, biological pleiotropy where one gene has two independent effects on traits, or spurious pleiotropy where two different genes affect each of the two traits^42^. The first or second might explain the *SHROOM3* effect, the latter might possibly include *MUC1/MTX1* linked to ADTKD^43^. It is noteworthy that the strongest representatives of the two distinct classes of filtration function only (“real eGFR only”) versus albumin leakage into urine only (“real UACR only”), mapping to *UMOD* or *CUBN*, respectively, are without any effects on the other trait. Thus, it is not typical that effects on filtration function reflect on albuminuria or vice versa. Our results provide important new insights into a new class of kidney function genetics with concordant effects on eGFR and UACR and underscore that this is a physiologically compelling group of variants that might have been underacknowledged so far.

Compared to previous single kidney trait GWAS^7–9,11^, our multi-trait GWAS identified novel associated regions including two signals, where index variants resided near genes known for renal cancer^17,18^ (*LOC440040* and *ZFHX4*). Among known kidney GWAS signals, eight genes were mapped by multi-trait high-PIP variants with >50% probability to be causal for the association signal that were missed by single-trait fine-mapping. These include previously described genes^11,23,44^ (*SLC34A1, OAF* and *EEF1AKMT2*) and novel likely causal genes for eGFR or UACR with pronounced expression in kidney cells (*RERG, APH1B, POR, MDH2* and *TMEM120A). RERG* encodes the RAS-like estrogen-regulated growth inhibitor^45^. *APH1B* is essential for Notch signalling, active in kidney development, silenced in healthy adult kidneys^46^, and contributing to proteinuria, diabetic kidney disease, and fibrosis when re-activated^47,48^. *POR*, *MDH2*, and *TMEM120A* all mapped to the same signal, which was associated with filtration function and, interestingly, with UACR but without evidence of albumin leakage into urine. The lead variant mapped to *POR* (missense) as well as *MDH2* and *TMEM120A* (kidney eQTLs), with its effect on UACR largely attributable to Ucrea changes driven by fluid intake through *POR*’s role in CYP activity and caffeine metabolism^49^. By contrast, *MDH2* and *TMEM120A* are more likely involved in kidney water handling via ATP-dependent sodium resorption and AQP2-mediated water resorption, respectively^31,32,50^.

When classifying associated signals as associations with eGFR, we wanted to make sure that variant associations were not purely creatinine or cystatin biomarker metabolism by requiring concordant associations on both eGFRcrea and eGFRcys as done previously^51^. We also wanted to ascertain that associations with UACR were not mere associations with Ucrea, the denominator in UACR, without any evidence for leakage into urine ^11^. We found that many of these “shadow UACR” effects were – at least in part – driven by fluid intake, particularly for the signals near *CYP1A1*, *POR*, and *AHR*, known for increased caffeine preference^49^ which appeared to also increase total fluid intake.

A key strength of our analysis is the multi-trait approach, which helps identify and fine-map signals when more than one trait association is involved (“real eGFR” or “both”). A general challenge in multi-trait studies is the comparison of GWAS results across traits at the level of association signals. For single traits, a widely used method for identifying independent association signals is approximate conditional analysis using GCTA^19^. However, this method is not directly applicable to multiple traits. To address this, we developed a novel workflow, MT-GCTA, which integrates GCTA into the context of multi-trait GWAS. A strength was also to incorporate BUN in the multi-trait GWAS, since its correlation with eGFRcrea and eGFRcys can help strengthen the association evidence. However, we did not involve BUN in the signal classification: of the signals robustly identified with eGFRcrea and eGFRcys association, only 94.3% aligned with directional concordant significant association with BUN, which would have de-classified numerous “real eGFR” signals. Regarding the claim for mapped genes, we have been rather strict, by requiring a probability for being causal of at least 50%, which might have missed interesting genes, but ascertained less false positives. Some limitations should be noted. Sample size and statistical power varied across traits, and “real UACR only” and “both” group signals may benefit from larger sample sizes for UACR. Additionally, Ualb and Ucrea were available only in UKB and measured as concentrations rather than as excretion rates, limiting interpretability in the absence of urine volume data. Furthermore, low Ualb was set to the (high) LOD in UKB, thus in a large proportion of the (kidney-healthy) individuals, the UACR associations are likely driven by Ucrea (because the nominator is fixed in these individuals). Regarding fluid intake, the GWAS was based on a simplified phenotype derived from aggregated intake of water, tea, and coffee assessed via frequency questionnaires. This enriched for caffeine preference, but using only water intake would have misrepresented general population habits. This also ignored fluid intake from other sources like soft drinks or diet. While we are aware that our fluid intake assessment is subject to high measurement error, the detection of genetic associations, gene expression enrichment in brain and overlap with associations on urinary creatinine concentration supported the phenotype as meaningful. Overall, measurement error in any variable, particularly Ualb or fluid intake, can lead to missed or underestimated associations, but would rather not result in false positives.

It is important to note that our findings were made in a population of rather healthy adults. While it has been previously shown that associations with quantitative traits in healthy also link to associations with disease traits (e.g. eGFRcrea and CKD^52^; BMI and obesity^53^), the impact of the “both” group signals on disease traits or in pathological status remains to be shown. An important question will be how concordant effects on eGFR and UACR in healthy translate to CKD patients and risk of kidney failure.

In summary, multi-trait GWAS and fine-mapping can successfully improve signal detection and resolution, which was key to identify associations with both filtration function and level of albuminuria and underlying mapped genes. We identified a new class of kidney function genetics with concordant effects on both eGFR and UACR with physiologically compelling mechanisms. Our genes mapped by highly likely causal variants and biological follow-up provide interesting new starting points for functional follow-up of the genes’ roles in physiological and pathological states of the kidney.

## METHODS

### Meta-analyses summary statistics for four kidney traits

Our multi-trait GWAS was based on European-only results from meta-analyses including GWAS from the CKDGen Consortium and UK Biobank, for estimated glomerular filtration rate based on serum creatinine (eGFRcrea), estimated glomerular filtration rate based on cystatin C (eGFRcys), blood urea nitrogen (BUN) and urinary albumin-to-creatinine ratio (UACR). All except BUN were publicly available^8,11^. In the published work, the BUN meta-analysis includes a subset of non-European individuals. We thus repeated the BUN meta-analysis according to the methods from the published work but excluding any non-European individuals.

We here used the European-ancestry results including 1,004,040 persons for eGFRcrea, 460,826 for eGFRcys, 679,531 for BUN and 547,361 for UACR. In brief, all GWAS were adjusted for age, sex and principal components and all traits were log-transformed, in addition UACR was inverse-normal transformed. Details on the outcome transformations can be found in the respective publications. Quality control on these data was performed on all four sets of GWAMA summary statistics. Using EasyQC^54^, multiple filtering steps were executed. Excluded were variants with low quality, i.e., sample size of less than 10,000, rare variants with effect allele frequencies below 1%. Additionally, all samples with missing or invalid values, all non-biallelic SNPs (as they were not analyzed in CKDGen for eGFRcrea) and all duplicates were removed. Furthermore, GC lambdas were calculated to infer confounding by population stratification^55^.

### Multi-trait GWAS based on meta-analysis summary statistics

The multi-trait GWAS method used was published in 2022 by Xiong et al.^14^ under the name C-GWAS. It has been described as more powerful than MTAG^15^ and other similar methods as well as being less sensitive towards different genomic backgrounds of input statistics, in part due to abandonment of LD-based calculations, which are part of most other tools^15,56^. This also increases computational efficiency, all the while maintaining similar rates of accuracy and estimation errors. Another benefit is that it only requires GWAS summary statistics as input^14,57^. In two steps, it calculates P-values for an association with any of the input traits and corrects for the multiple testing burden of multiple traits. The derived P-values can directly be compared to single-trait GWAS results with a genome-wide significance threshold^57^ (P<5×10^-8^). In our case, we only included SNPs that occur in the summary statistics of all four traits (eGFRcrea, eGFRcys, BUN and UACR). In total, 7,192,814 SNPs were analyzed. All effect alleles and corresponding estimates were turned to match the allele with the smaller allele frequency prior to the C-GWAS analysis.

### Definition of multi-trait GWAS regions

First, for each of the four kidney traits (eGFRcrea, eGFRcys, BUN and UACR) separately, genome-wide significant variants (P<5×10^-8^, Bonferroni-corrected for ∼1M independent tests genome-wide) were selected and combined into independent regions (requiring d>500kb between two genome-wide significant variants of two regions, as described in Stanzick et al.^8^. We then combined single-trait regions across the four kidney traits and aggregated overlapping regions into a new “multi-trait” (MT) region (again requiring d>500kb between two regions). The lead variant of each MT-region was determined as the variant with the highest sum of explained variance across all four traits. Explained variance for an individual trait was calculated as follows:

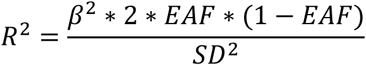

Standard deviations of *SD_eGFRcrea_* = 0.16, *SD_eGFRcys_* = 0.2, *SD_BUN_* = 0.24 and *SD_UACR_* = 1 were used (calculated from available UK Biobank phenotype data). The explained variances were summed to determine the MT-region lead variant, i.e., the variant with the highest sum of explained variance. If the association P-value did not reach the significance level of P<10^-5^, the explained variance for this trait was set to zero before adding it to the explained variances of the other traits. Region coordinates of regions only identified by C-GWAS were checked for overlap within a ± 500kb distance around the region with the position of independent signal or region lead variants of previously published data^7–11^.

### Multi-trait approximate conditional analyses using GCTA

Single-trait approximate conditional analysis using GCTA^19^ was expanded to be used with multiple traits, using the (maximum) explained variance of the SNP across traits instead of the (minimal) P-value to determine the signal index variant. Cleaned summary statistics of all four main traits, a list of multi-trait regions and their coordinates, and the standard deviation of each main trait was needed for this analysis. A detailed description and workflow can be seen in **Supplementary Note 1** and **Supplementary Figure 4**. In short, (i) the sum of explained variance across all four traits is calculated for each SNP in the multi-trait region, (ii) the first signal index variant is the SNP with the highest explained variance across all traits and all other SNP estimates are conditioned on this SNP using “--cojo-cond”, (iii) by checking for genome-wide significant conditioned P-values and recalculating the explained variance now based on conditioned estimates, further independent signal index variants are found, (iv) steps (ii) and (iii) are repeated until no further genome-wide significant SNPs are found. Lastly, fully conditioned effect estimates are calculated using “--cojo-joint” for all independent signal index variants.

### Multi-trait fine-mapping

All significant independent multi-trait signals underwent single-trait fine mapping and multi-trait fine-mapping. First, the traditional single trait fine-mapping approach was applied, for which we derived 95% credible sets based on approximate Bayes factors that were derived using the Wakefield formula^20^. The Wakefield results were then transferred as input for multi-trait fine-mapping using flashfm^13^ which requires single trait fine-mapping results as input. As described by the authors, only traits that have at least one variant with a P-value <10^-6^ in the signal were added to the analysis, and at least two traits were required to fulfil this criterion for a signal to be fine-mapped by flashfm. In signals that did not have more than one trait that fulfilled this criterium, flashfm fine-mapping was not performed. After correspondence with the programmer, updated flashfm functions published as part of flashfmZero were used for the final analysis^58^. Flashfm does not find new signals, it recalculates the credible sets and posterior inclusion probabilities (PIPs) for already existing signals and for each trait separately, hence the need for single-trait fine-mapping statistics as input. Single-trait and multi-trait fine mapping results were compared using the total number of credible sets with specific sizes as well as the number of sets that contained a variant with a high-PIP (PIP>0.5).

### Variant annotation and gene mapping

High-PIP (PIP>0.5) variants were annotated for functional consequences on gene products using annotation data from dbSNP version 151^21^ and annotated for their regulatory effects on gene expression in two kidney tissues (glomerular and tubulo-interstitial tissues) from NephQTL2^22^. Significant effects on expression were selected at FDR<5%.

### UK Biobank GWAS for primary and secondary traits

For further interpretation purposes, summary statistics of UKB data were used for the traits eGFRcrea, eGFRcys, BUN, UACR, urinary albumin (Ualb) and urinary creatinine (Ucrea). eGFRcrea and eGFRcys were measured in ml/min/1.73², winsorised at 15 or 200ml/min/1.73², and log transformed. Blood urea was measured in mmol/l, multiplied by 2.8 to generate BUN values and log transformed. Ucrea was measured in mmol/l and multiplied by 88.4 to change the unit to mg/l and Ualb was measured in mg/l and values below 6.7 were set to 6.7mg/dl. UACR values were log and inverse-normal transformed and adjusted for age and sex. For Ucrea and Ualb GWAS, the traits were transformed as described for UACR. The advantage of limiting the data to UKB for comparison between traits is to then have a comparable sample size of approximately N∼440,000 participants between all traits. Additionally, GWAS summary statistics were calculated dichotomized eGFR<60ml/min/1.73² (yes/no) and dichotomized UACR≥30mg/g (yes/no) following the definition of the KDIGO CKD Working Group ^5^. All UKBB GWAS were adjusted for age, age^2^, age x sex, age^2^ x sex, sex and the first 20 principal components. Age was mean centered to avoid multicollinearity. QC was performed like meta-analysis QC, with the exception that the effect allele frequency filter was removed, since some variants would be excluded that would pass this filter in the meta-analysis results. To derive fully conditioned effect estimates (conditioned on other signals in an associated region) for all traits, we used GCTA to condition each trait summary statistics on the multi-trait signals index variants that were identified by using meta-analysis data using “--cojo-joint”. Signals identified by the multi-trait GWAS based on meta-analyses were tested for their association in UKB using a threshold adjusted for multiple testing (Bonferroni-correction on the number of multi-trait signals). Additionally, mean arterial pressure (MAP) and haemoglobin A1c (HbA1c) were analysed as secondary traits. MAP was calculated with the following formula from UKB data: MAP = DBP + (SBP-DBP)/3 [mmHg], with SBP and DBP being systolic and diastolic blood pressure, respectively. No further transformation was applied due to normal distribution of MAP. HbA1c percentages were calculated from measured HbA1c [mmol/mol] by HbA1c_percentage = (HbA1c [mmol/mol] / 10.929) + 2.15. The HbA1c percentages were inverse-normal transformed to obtain normality.

### Classification for “real eGFR” and “real UACR”

UKB associated signals were tested for association with eGFRcrea and eGFRcys to determine ”real eGFR” associated signals using fully conditioned effect estimates (expecting concordant effect direction and statistically significant eGFRcrea and eGFRcys association, using Bonferroni-correction on number of signals, P_c_<0.05/740). Similarly, signals were tested for their association with UACR (using Bonferroni-correction, P_c_<0.05/740) to determine “real UACR” associated signals. To ensure a UACR association was driven by Ualb, the association of UACR variants was further tested for increased Ualb effects (using Bonferroni-correction on the number of UACR signals, P_c_<0.05/78). The obtained signals were classified based on their “real eGFR” and “real UACR” effect directions. The signals not classified as “real UACR” were then tested for their association with Ucrea (Ucrea P_c_<0.05/78, Ualb P_c_>0.05/78) to determine Ucrea driven UACR associations (“shadow UACR”). Effect directions were generally adjusted to eGFR-decreasing (i.e., filtration function decreasing) alleles, except for variants that were only associated with UACR (and not “real eGFR”), for which alleles were adjusted to UACR-increasing alleles. All effect sizes were standardized (dividing by outcome SD) to allow for comparison of effect sizes between traits.

### Kidney cell-type specific expression

Mapped genes were queried for their cell-type specific expression in kidney cell-type specific expression data^26^ and in data from the kidney cell atlas^25^ (KPMP). We counted the number of genes that were differentially expressed in at least one cell-type of a kidney substructure and compared the relative frequency of genes per substructure between classes.

### Fluid intake GWAS in UK Biobank and tissue enrichment

Fluid intake was calculated as the sum of cups drunken per day of water, coffee and tea and transformed into liter per day from UKB data. All three fluids were measured in self-reported cups drunken per day and included decaffeinated coffee and black and green tea^59–61^. Other fluids such as juice, soft drinks and alcoholic beverages were not reported at the same time as the other three. GWAS results were calculated, and fully conditioned effect estimates were generated as described for other UKB summary statistics above. Additionally, the fluid intake GWAS was evaluated to derive genome-wide significant (P<5.0×10^-8^) associated signals using a distance- and LD-based clumping criterion (d<500kb, r2>0.01). GTEx tissue expression enrichment analysis was conducted based on the genome-wide fluid intake summary statistics applied to FUMA-MAGMA^62^. UKB GWAS results on each contributor of our aggregated fluid intake variable, i.e., on water, coffee and tea intake separately, were queried using data from GWAS atlas^63^.

### Mediator analyses

We conducted causal mediator analyses using the mediate() function of the R package mediation^34^. Based on the Baron-Kenny method^64^ and linear regression on the mediator and the outcome (adjusted for the mediator), the package infers direct (mediator-adjusted effects) and indirect (effects through the mediator) effects of the genetic variants on an outcome. Mediator analyses were conducted for several combinations of primary and secondary traits using individual-participant data of unrelated European individuals from UKB (N=356,000). All outcomes were inverse-normal transformed before mediator analyses to obtain comparable effect sizes.

### Correlation analyses

Phenotypic correlations between traits were estimated using Spearman correlation based on age and sex adjusted UKB phenotype data. Genetic correlation between traits were estimated using LD-score regression applied to the UKB GWAS summary statistics^65^.

### Associations with other traits and enrichment tests

Several subsets of signals were evaluated for their effects on Ucrea, fluid intake, MAP, HbA1c. Enrichment analyses were conducted using binomial tests using binom.test() in R. The enrichment tests were conducted as one-sided tests against Bonferroni-corrected P-values per class (P<0.05/number of signals per class).

## Data Availability

The raw datasets supporting the conclusions of this article can be applied for from UK Biobank (https://www.ukbiobank.ac.uk/). C-GWAS and meta-analyses summary statistics are available online for eGFRcrea and eGFRcys from https://www.genepi-regensburg.de/gwas-summary-statistics, and for UACR from https://ckdgen.imbi.uni-freiburg.de/datasets/Teumer_2019.

## DECLARATIONS

### Ethics approval and consent to participate

Not applicable.

### Consent for publication

Not applicable

### Data and code availability

The code to conduct multi trait GCTA conditioning is available from https://github.com/genepi-regensburg/mtgcta. All other code is available from the corresponding author on reasonable request. The raw datasets supporting the conclusions of this article can be applied for from UK Biobank (https://www.ukbiobank.ac.uk/). C-GWAS and meta-analyses summary statistics are available online for eGFRcrea and eGFRcys from https://www.genepi-regensburg.de/gwas-summary-statistics, and for UACR from https://ckdgen.imbi.uni-freiburg.de/datasets/Teumer_2019.

### Author contributions

T.W.W. conceived the experiments, supervised the project, conducted mediation analyses and contributed to the writing group. H.C.d.H. conducted all main analyses, wrote the first manuscript draft and contributed to the writing group. I.M.H. supervised the project and contributed to the writing group. B.F.G. conducted cell-type specific expression analyses. A.T. contributed to the development of the UACR classification. K.J.S. and R.W. contributed to the interpretation of biological follow up. All authors reviewed the manuscript.

