## Supplementary Material for "Distinct and shared genetics of kidney filtration function versus albuminuria revealed by multi-trait GWAS"

**SUPPLEMENTARY TABLES**

**Supplementary Table 9: Nine “shadow UACR” variants associated with fluid intake.**

Shown are 9 signal index variants that were associated with “shadow UACR” (with UACR, N=444,861; and with Ucrea, N=444,861) and fluid intake (N= 453,221, P_c_<0.05/31, corrected for 31 “shadow UACR” variants) in UKB. Effect estimates are fully conditioned (Ualb from UKB, N=444,861). Bonferroni-corrected significant associations are marked in bold (Pc<0.05/740 for eGFRcrea, eGFRcys and UACR, Pc<0.05/78 for Ucrea). All signals are not associated with Ualb (P_c_>0.05).

|  |  |  |  |  | **eGFRcrea** | | **eGFRcys** | | **UACR** | | **Urinary creatinine** | | **Fluid intake** | |
| --- | --- | --- | --- | --- | --- | --- | --- | --- | --- | --- | --- | --- | --- | --- |
| **RSID** | **Nearest Gene** | **OA** | **EA** | **EAF** | **beta** | **p** | **beta** | **p** | **beta** | **p** | **beta** | **p** | **beta** | **p** |
| rs2472297 | *CYP1A1* | c | t | 0.26 | 0.0031 | **1.1E-17** | 0.0024 | **4.3E-8** | 0.028 | **1.4E-30** | -0.03 | **5.5E-35** | 0.051 | **6.9E-170** |
| rs4410790 | *AHR* | t | c | 0.63 | 0.0028 | **8.2E-18** | 0.0015 | 2.1E-4 | 0.026 | **3.3E-33** | -0.022 | **2.9E-23** | 0.04 | **7.9E-124** |
| rs1057868 | *POR* | c | t | 0.28 | 0.0033 | **6.4E-21** | 0.0017 | **4.5E-5** | 0.013 | **9.1E-8** | -0.015 | **3.5E-10** | 0.024 | **3.6E-41** |
| rs2793351 | *DNAJC1* | g | a | 0.69 | -0.00089 | 0.0095 | 0.00053 | 0.19 | 0.014 | **4.8E-10** | -0.015 | **1.6E-10** | 0.0093 | **9.0E-8** |
| rs2068888 | *CYP26A1* | a | g | 0.55 | 0.0035 | **1.2E-28** | 0.0035 | **3.5E-20** | 0.014 | **1.4E-10** | -0.01 | **9.8E-7** | 0.0086 | **1.3E-7** |
| rs79600176 | *CYP2A7* | c | t | 0.98 | 0.0037 | 6.6E-4 | 0.0012 | 0.35 | 0.036 | **8.4E-7** | -0.039 | **1.0E-7** | 0.021 | **1.8E-4** |
| rs1936806 | *RSPO3* | t | c | 0.55 | 0.00015 | 0.63 | -0.0014 | 0.0025 | 0.0088 | **3.9E-5** | -0.0076 | **3.4E-4** | 0.0059 | **3.0E-4** |
| rs35153 | *CDH11* | a | g | 0.53 | 0.00095 | 0.0028 | 0.00062 | 0.11 | 0.0085 | **6.3E-5** | -0.0085 | **6.5E-5** | 0.0052 | **0.0013** |
| rs78444298 | *EDEM3* | a | g | 0.98 | 0.013 | **1.6E-29** | 0.008 | **8.5E-9** | 0.044 | **1.5E-8** | -0.042 | **4.9E-8** | 0.019 | **0.0013** |

**SUPPLEMENTARY FIGURES**

**Supplementary Figure 1. Workflow**

Shown is the general workflow of the analyses performed. Based on European GWAS summary statistics from meta-analyses of UK Biobank (UKB) and CKDGen for four primary kidney traits (eGFRcrea, eGFRcys, BUN and UACR, N up to ~1M), we conducted a C-GWAS multi-trait analysis. We selected significant signals from the multi-trait GWAS as well as from the single trait GWAS (*Data from Stanzick et al, 2023, $Data from Teumer et al, 2019). We conducted multi-trait fine-mapping using flashfm and classified the resulting signals regarding different kidney biology based on enrichment and mediator analyses with additional related traits.

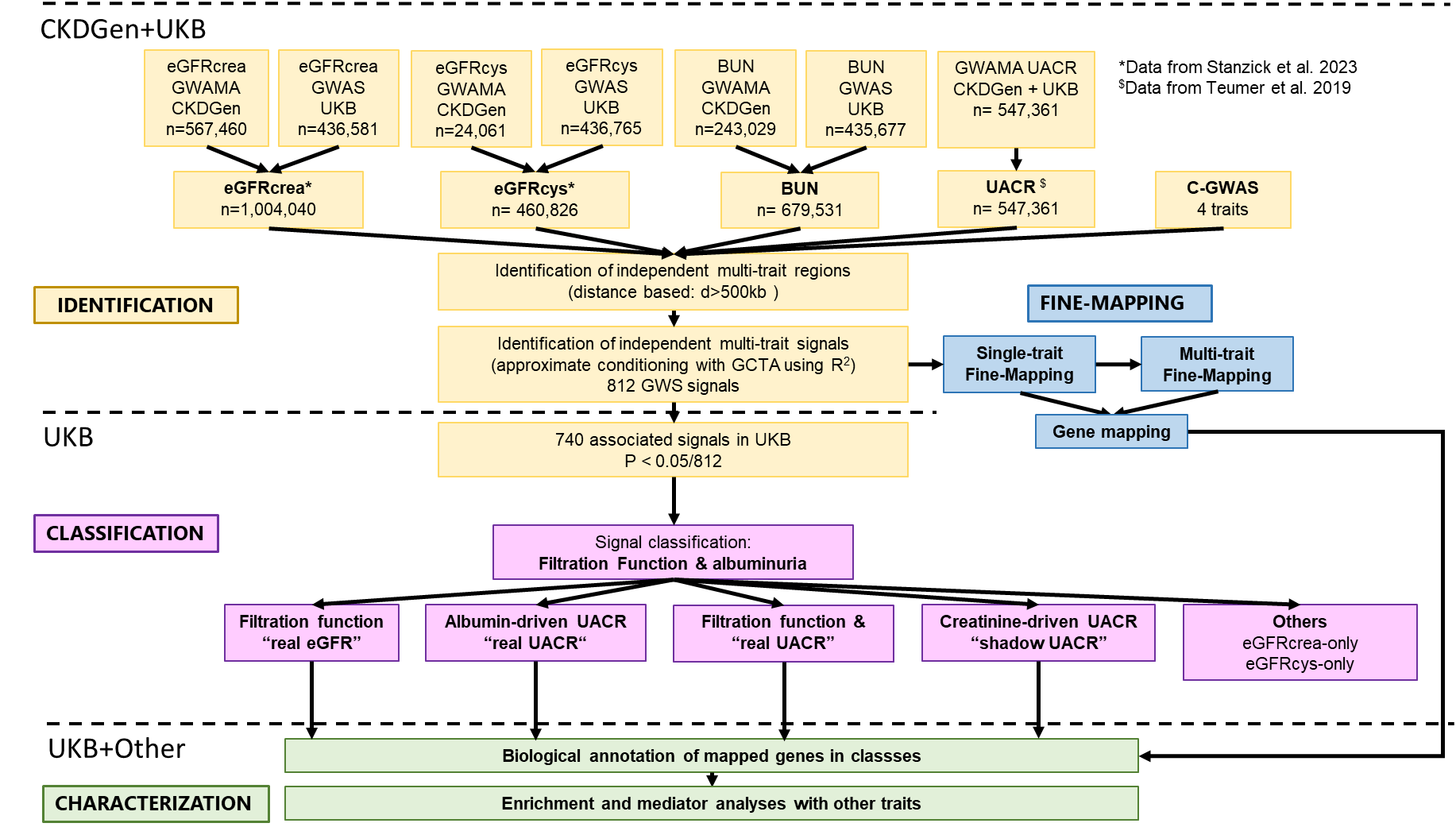

**Supplementary Figure 2. Single-trait GWAS association results**

Shown are the Manhattan and Quantile-Quantile plots for the meta-analysis results (CKDGen +UKB) and the multi-trait GWAS for **A**, **E**) eGFRcrea (N=1,004,040), **B**, **F**) eGFRcys (N=460,826), **C**, **G**) BUN (N=679,532), **D**, **H**) UACR (N=547,361). **A**-**D**) shows association P-values over chromosomal position. The QQ plots show the distribution of association P-Values based on all variants.

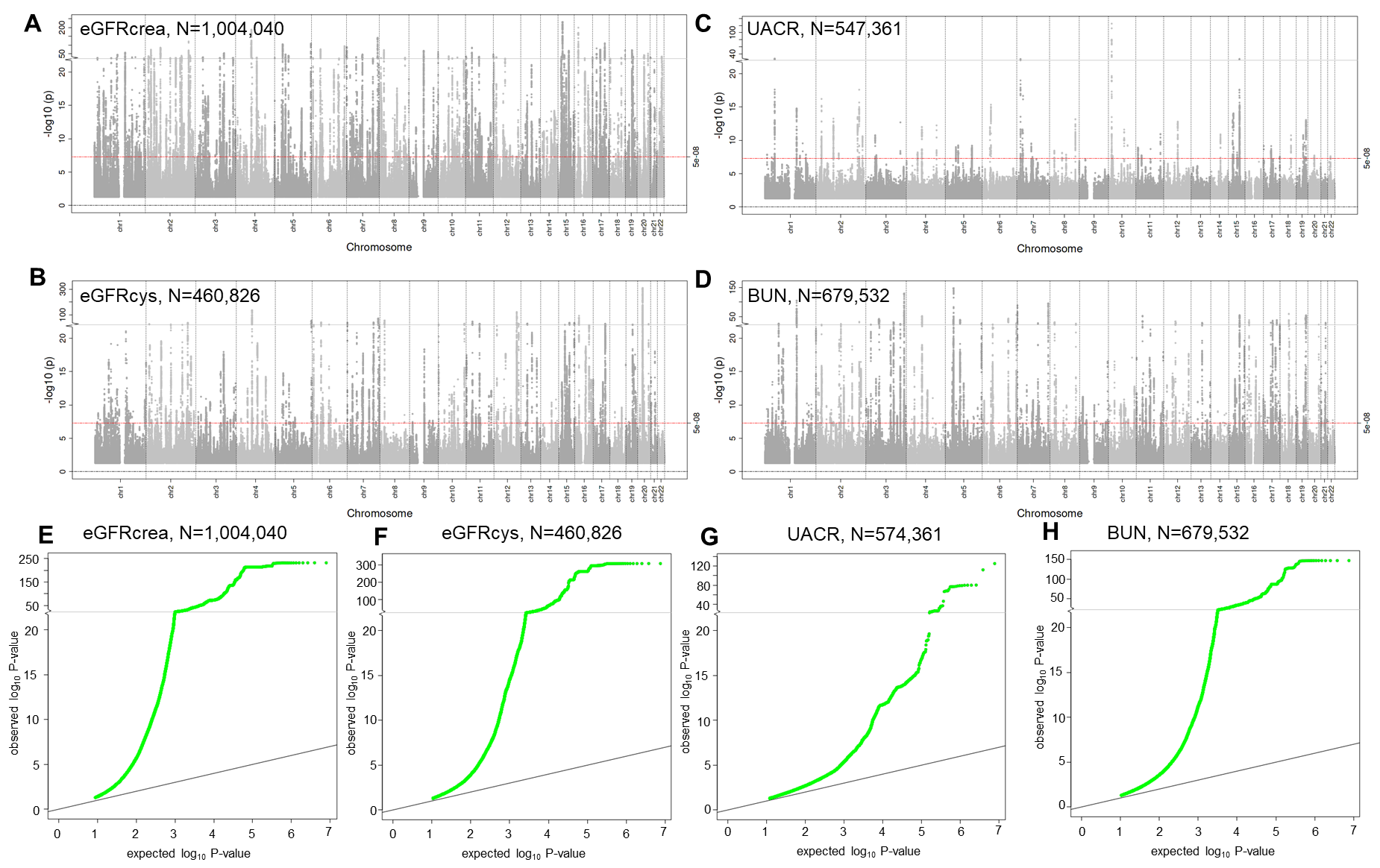

**Supplementary Figure 3. Region plots for novel kidney trait regions.**

Depicted are region plots unconditioned P-values for the 9 novel loci for CKDGen+UKB meta-analysis data. The depicted loci are named after the gene nearest the region lead variant. **A**) *TMEM31* (Region ID 468), **B**) *SENP7* (Region ID 560), **C**) *ELF2* (Region ID 567), **D**) *ZFHX4* (Region ID 486), **E**) *PTCSC2* (Region ID 571), **F**) *GRM5P1* (Region ID 303), **G**) *SCGB1A1* (Region ID 496), **H**) *AVPR1A* (Region ID 455), **I**) *LILRB5* (Region ID 575). Region plots were computed using LocusZoom standalone with a LD reference panel based on 20,000 unrelated Europeans from UK Biobank. Unconditioned p-values were used. Region plot axes: left y-axis: negative logarithmized association P-value, red line shows a genome-wide significance level of P<5x10^-8^, right y-axis: recombination rate in centimorgan per mega base (cM/Mb), x-axis: position on the chromosome in mega base (Mb).

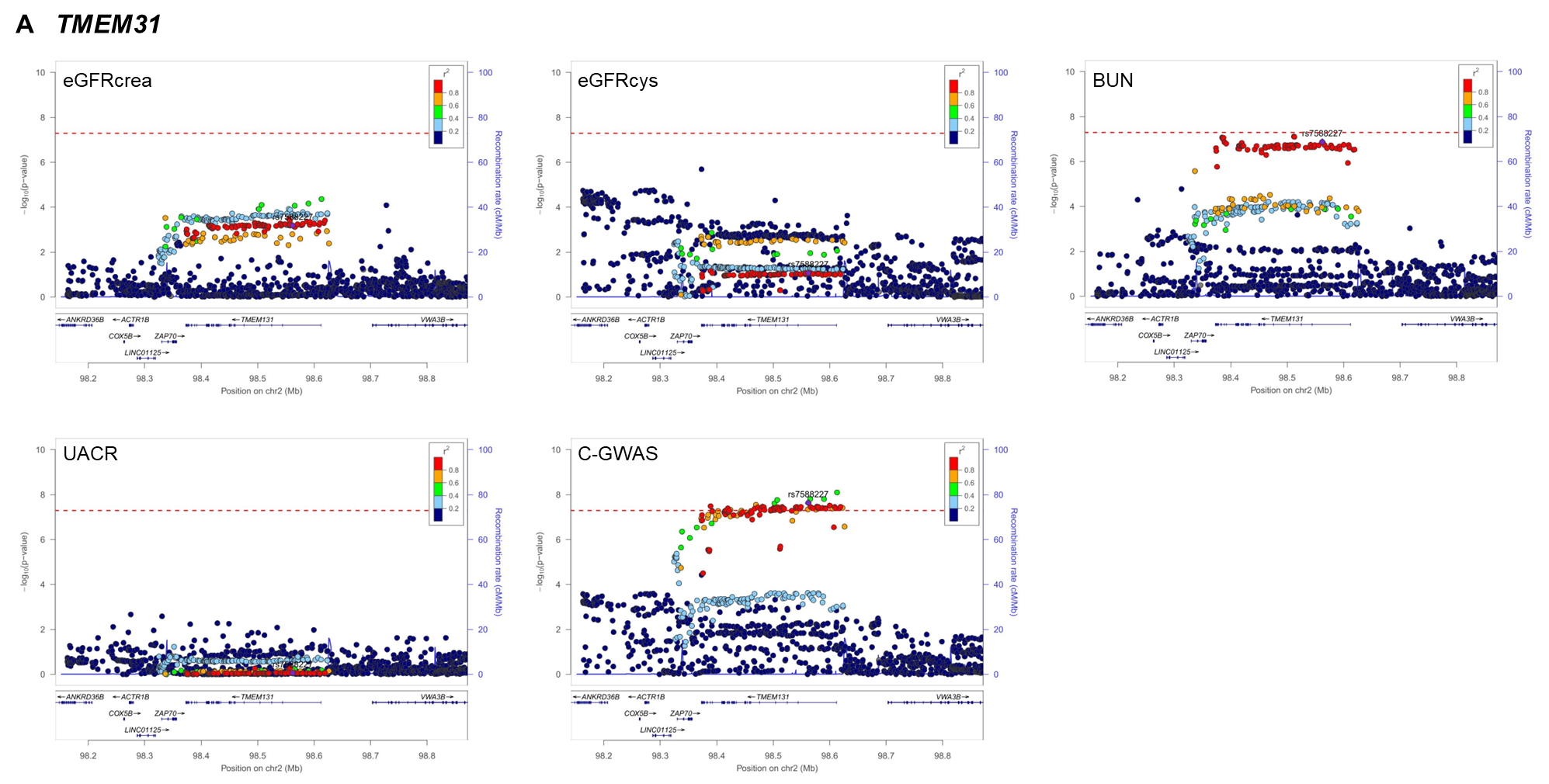

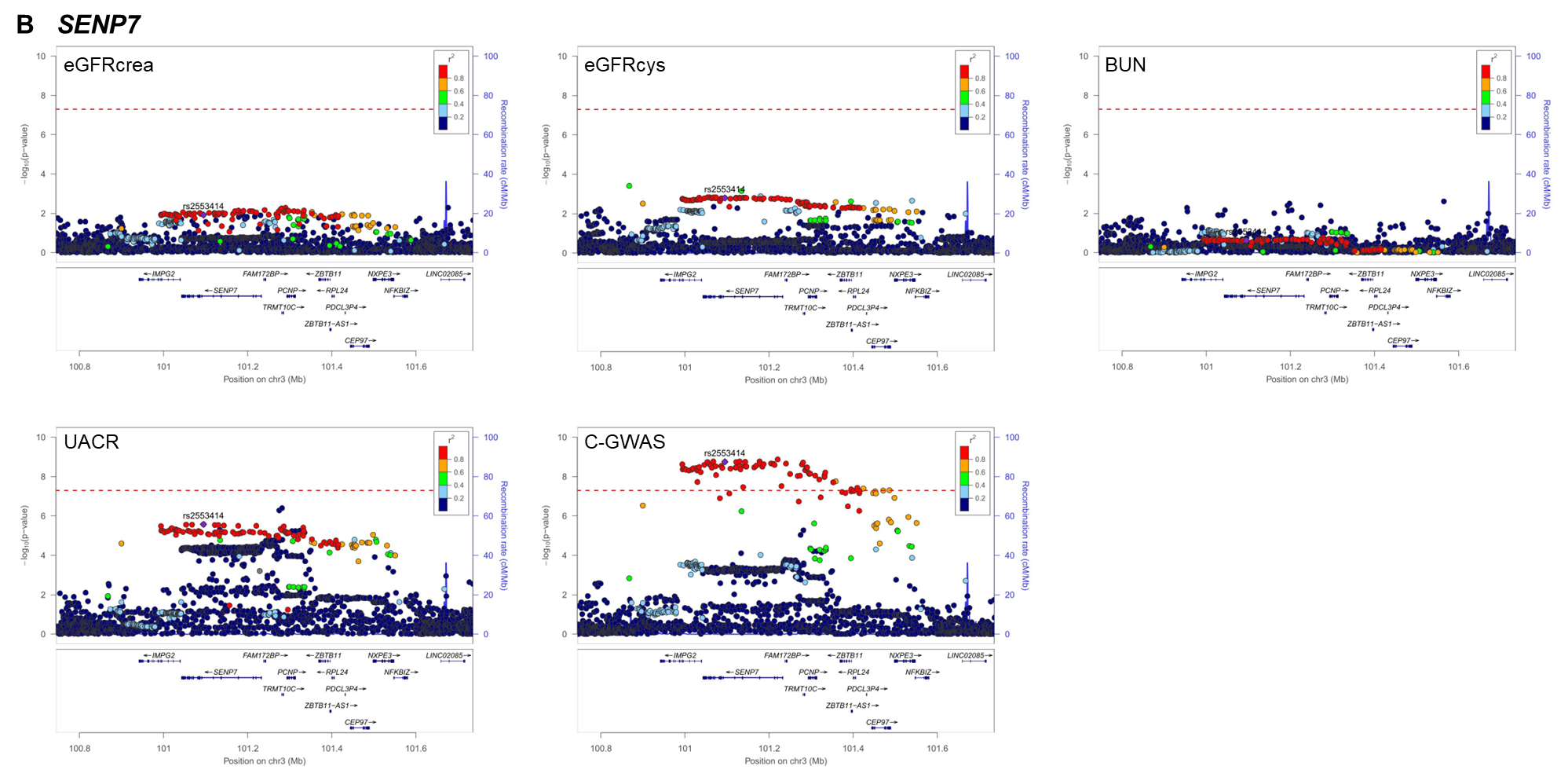

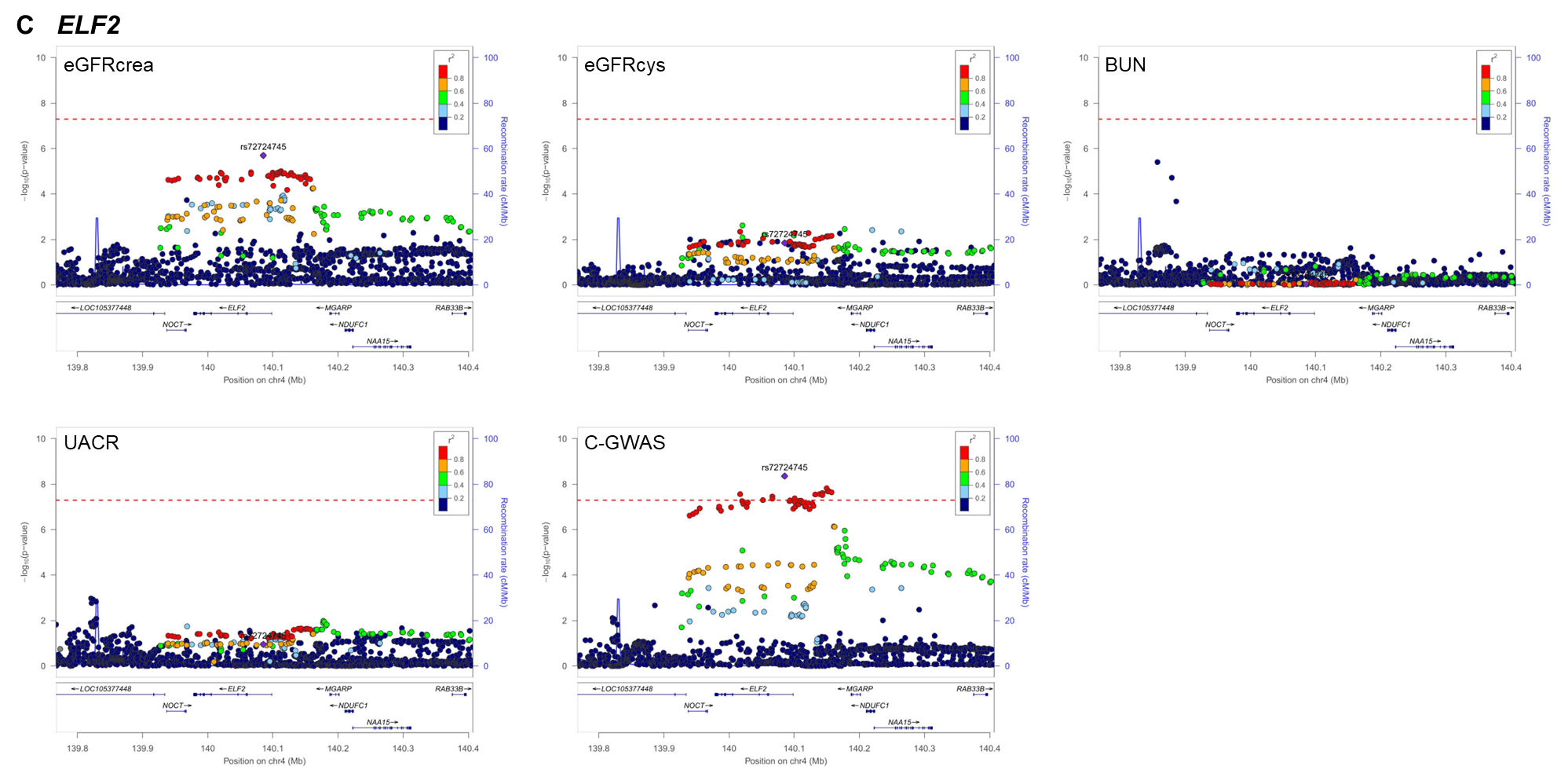

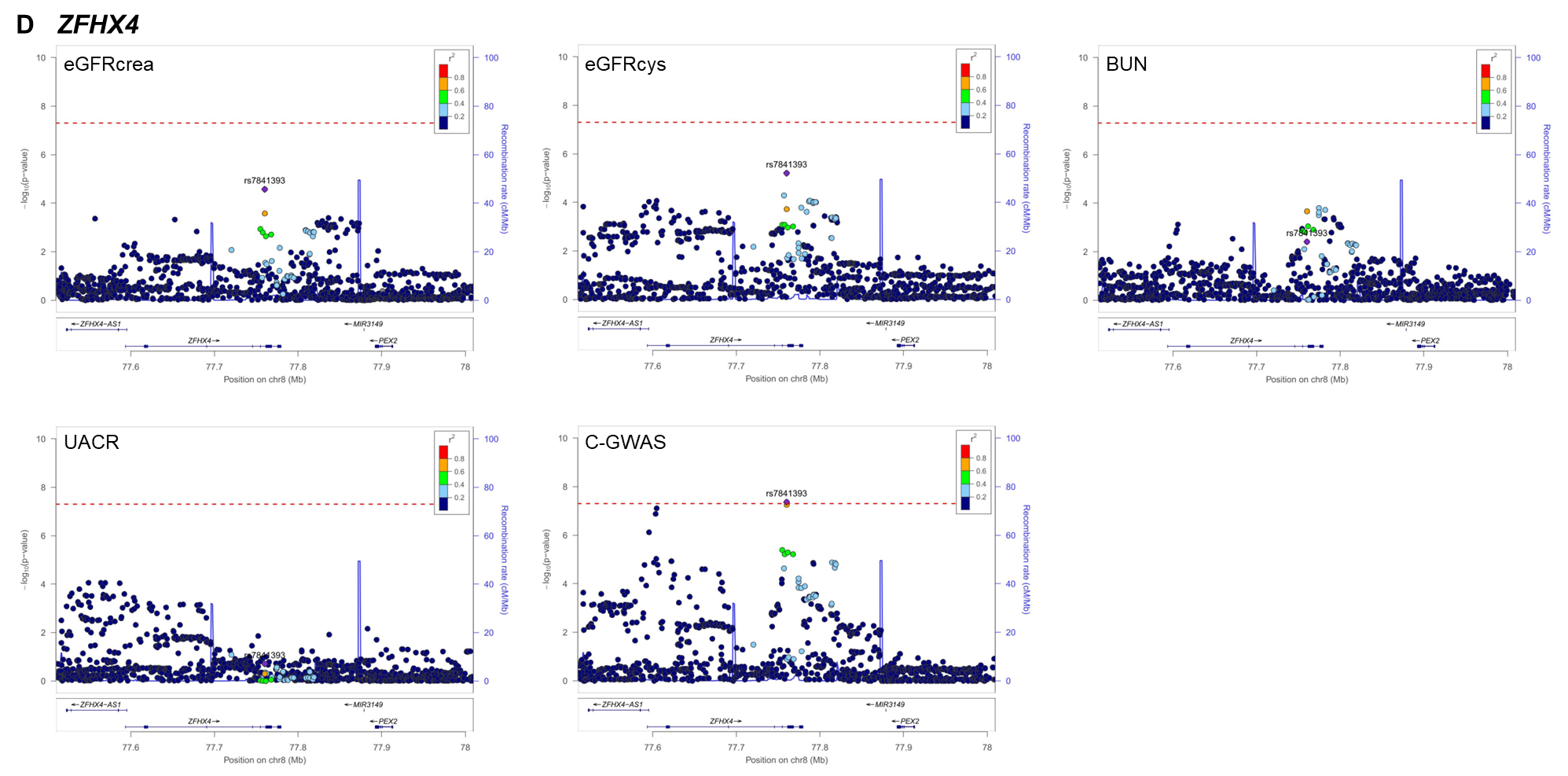

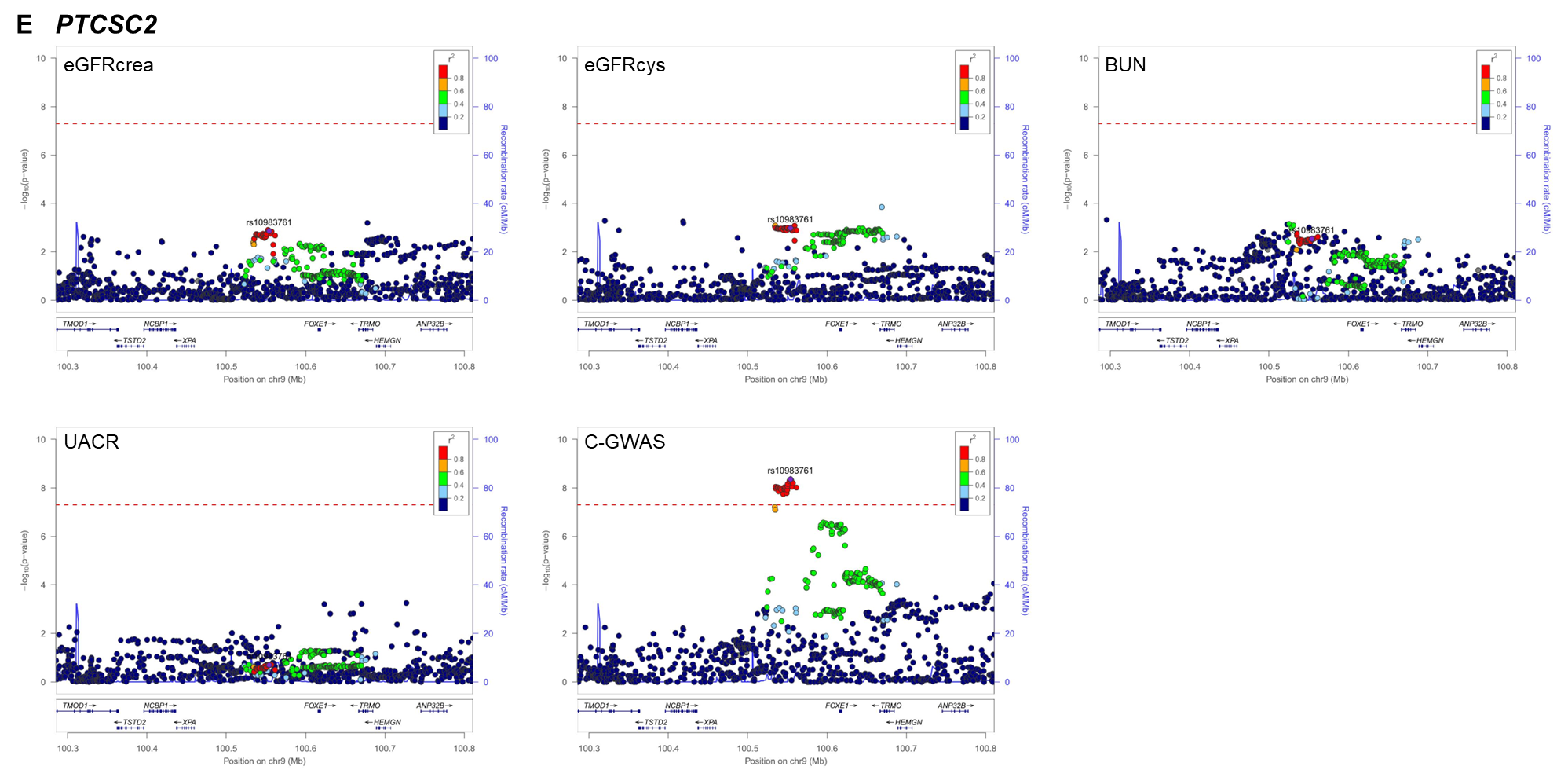

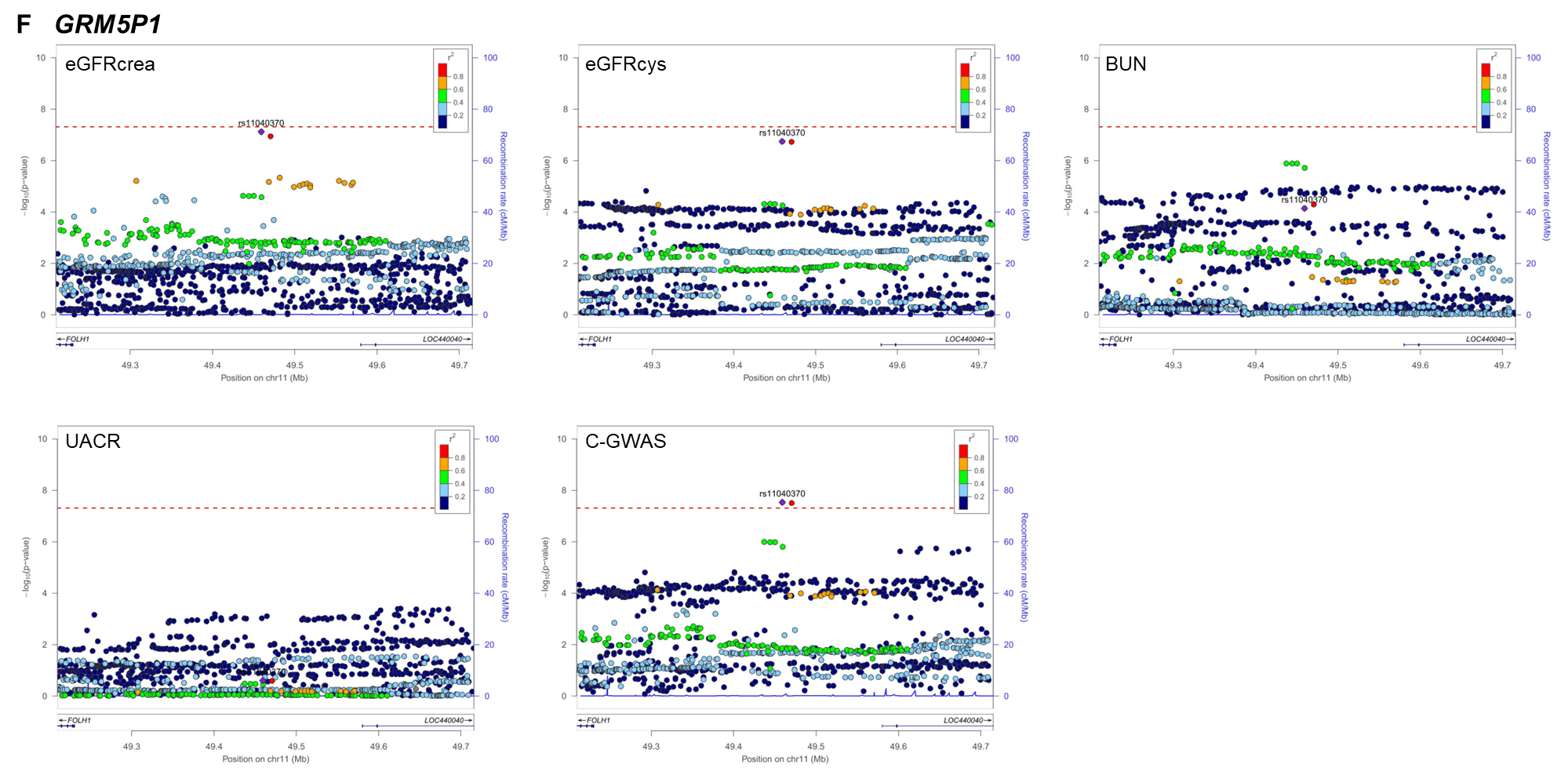

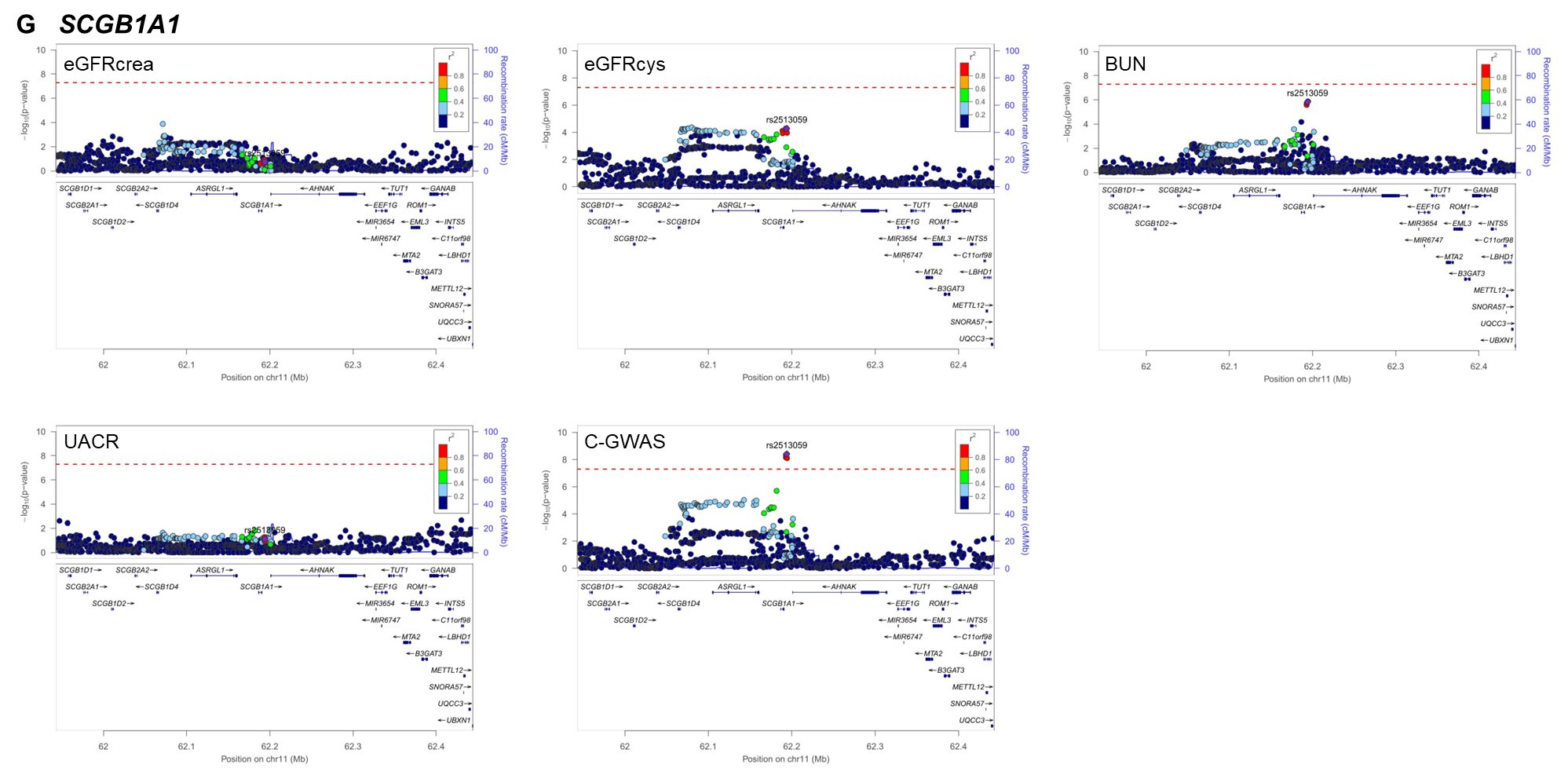

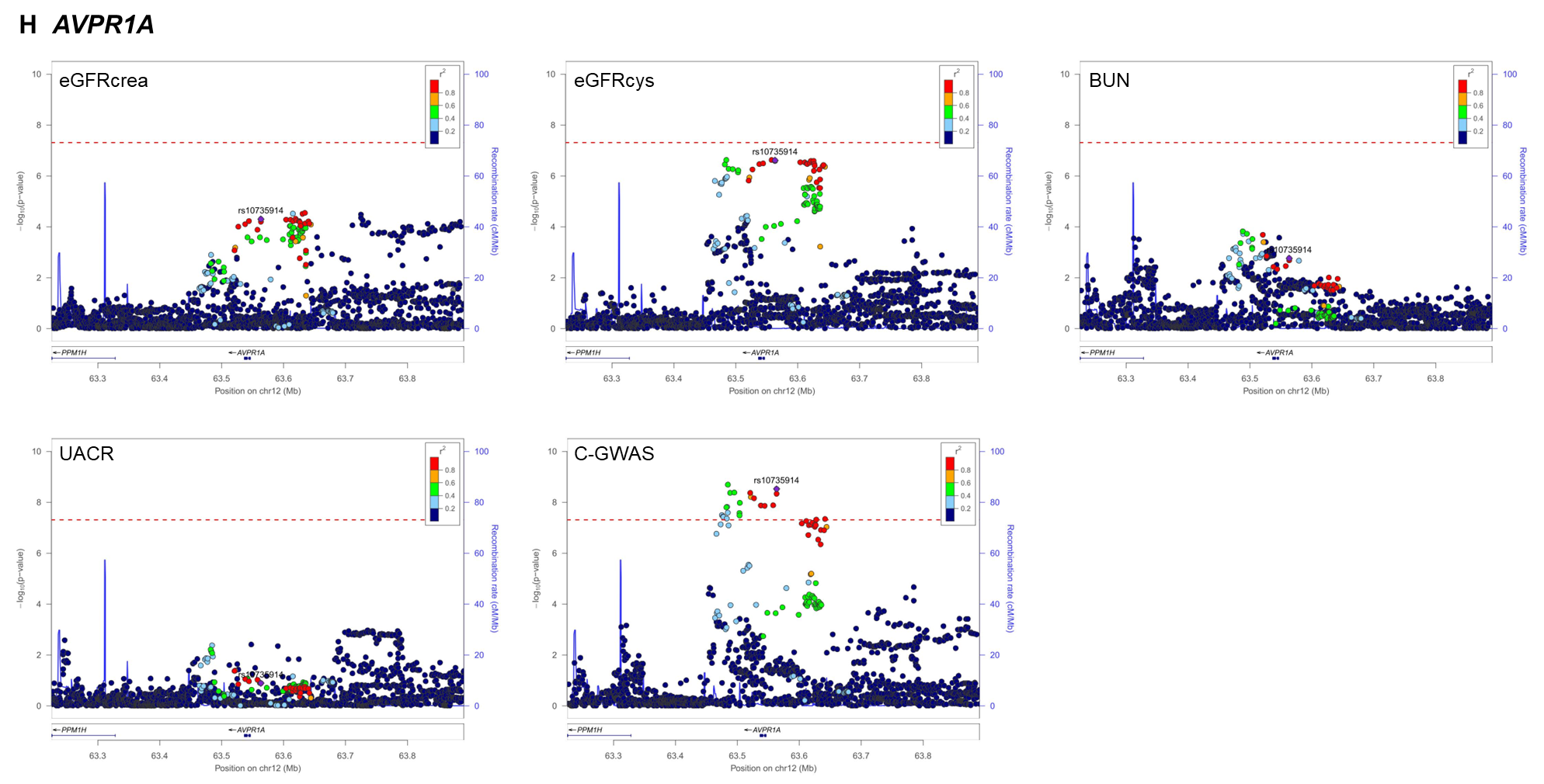

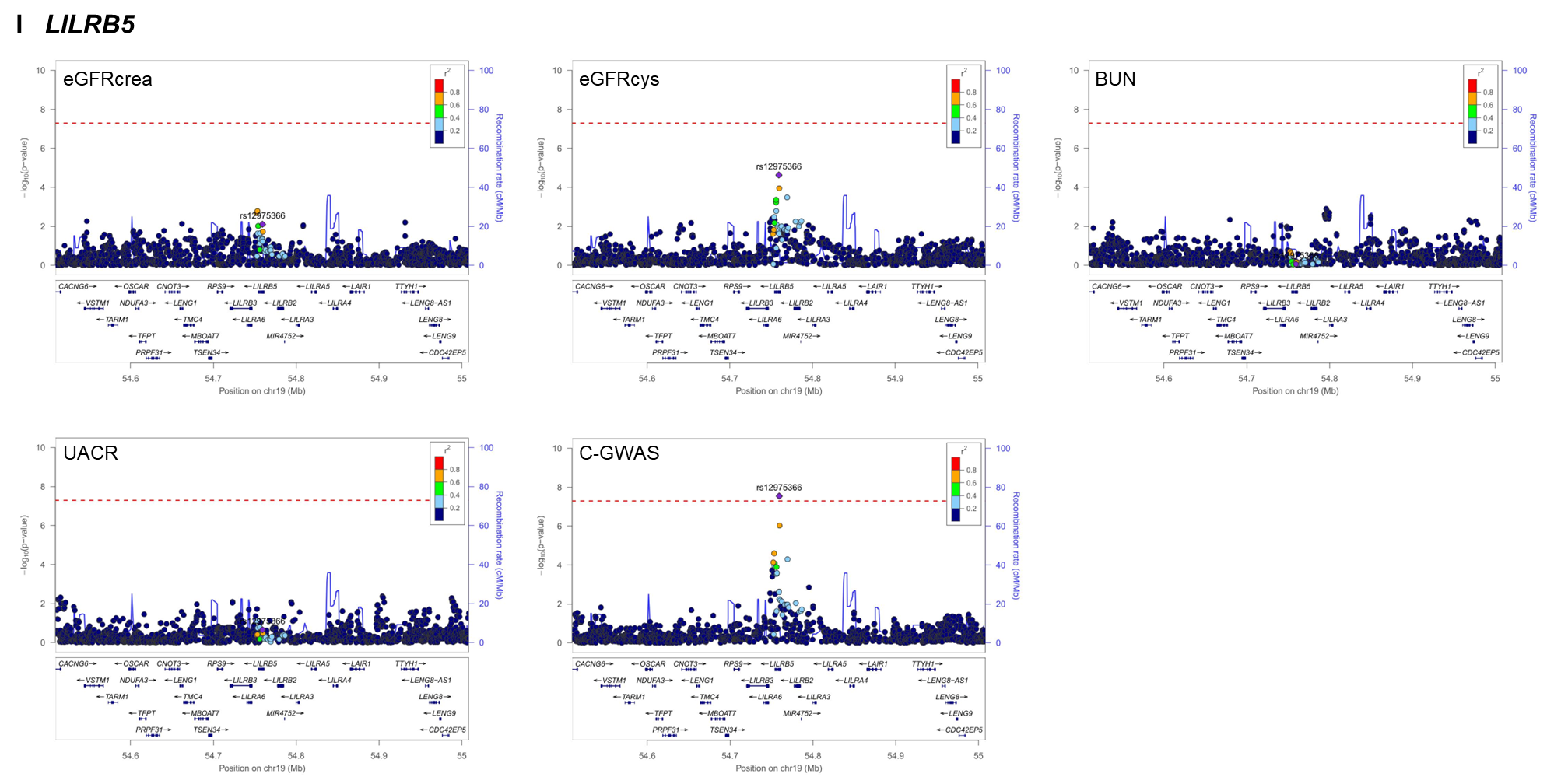

**Supplementary Figure 4. Algorithm of approximate conditioning with GCTA extended for multi-trait use**

The flow chart shows the process of multi-trait approximate conditional analysis with GCTA. In short, the sum of explained variance across all four traits is calculated for each SNP in the multi-trait region. If the association P-value does not reach P_c,i_<10^-5^, the explained variance for this variant in this trait was set to 0. The signal index variant is the variant with the highest sum of explained variance R² across all traits. All other SNPs are conditioned on the i signal index variants using “cojo-cond”. Conditioned P-values (P_c,i+1_) are checked for genome-wide significance and R_i_^2^ is recalculated on conditioned effect estimates. This process is repeated i-times until no further genome-wide significant P-values are found. Using “cojo-joint”, fully conditioned effect estimates are generated using the identified index variants are generated for each trait in each region. β_c,i+1,_ SE_c,i+1_: beta and standard error conditioned on i+1 signal index variants, SE_c,i+1._

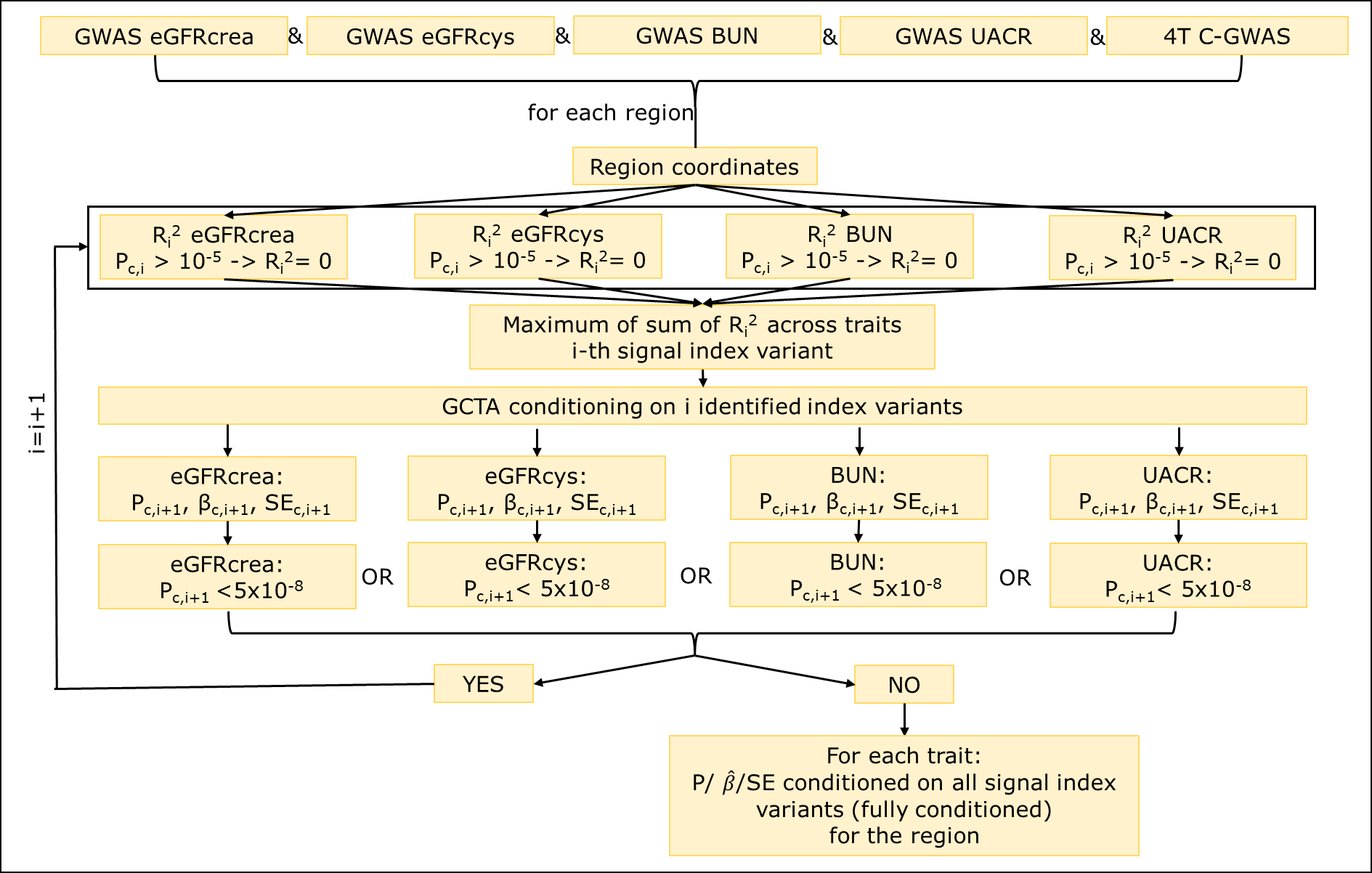

**Supplementary Figure 5: BUN association of eGFR associated signal index variants**

Signal index variants were tested for their association in UKB for the traits eGFRcrea (n=436,581), eGFRcys (n=436,765), BUN (n=435,677) and UACR (n=444,861). They were used for further analysis if they were associated with P_c_<0.05/812=6.16x10^-5^ with any of the four traits. Signal index variants were classified into eGFRcrea (P_c,eGFRcrea_<0.05/740, P_c,eGFRcys_ >0.05), eGFRcys (P_c,eGFRcys_<0.05/740, P_c,eGFRcrea_>0.05) and “real eGFR” (P_c,eGFRcrea_ & P_c,eGFRcys_<0.05/740, consistent effect direction). Additionally, they were tested for their association with BUN (P_BUN_<0.05/740, depicted by filled circles). Alleles were turned to match eGFR decreasing effect sizes. Depicted are BUN effect estimates in comparison to **A**: eGFRcrea and **B**: eGFRcys. In both cases a few “real eGFR” signals would not be classified as such, if BUN was used for classification.

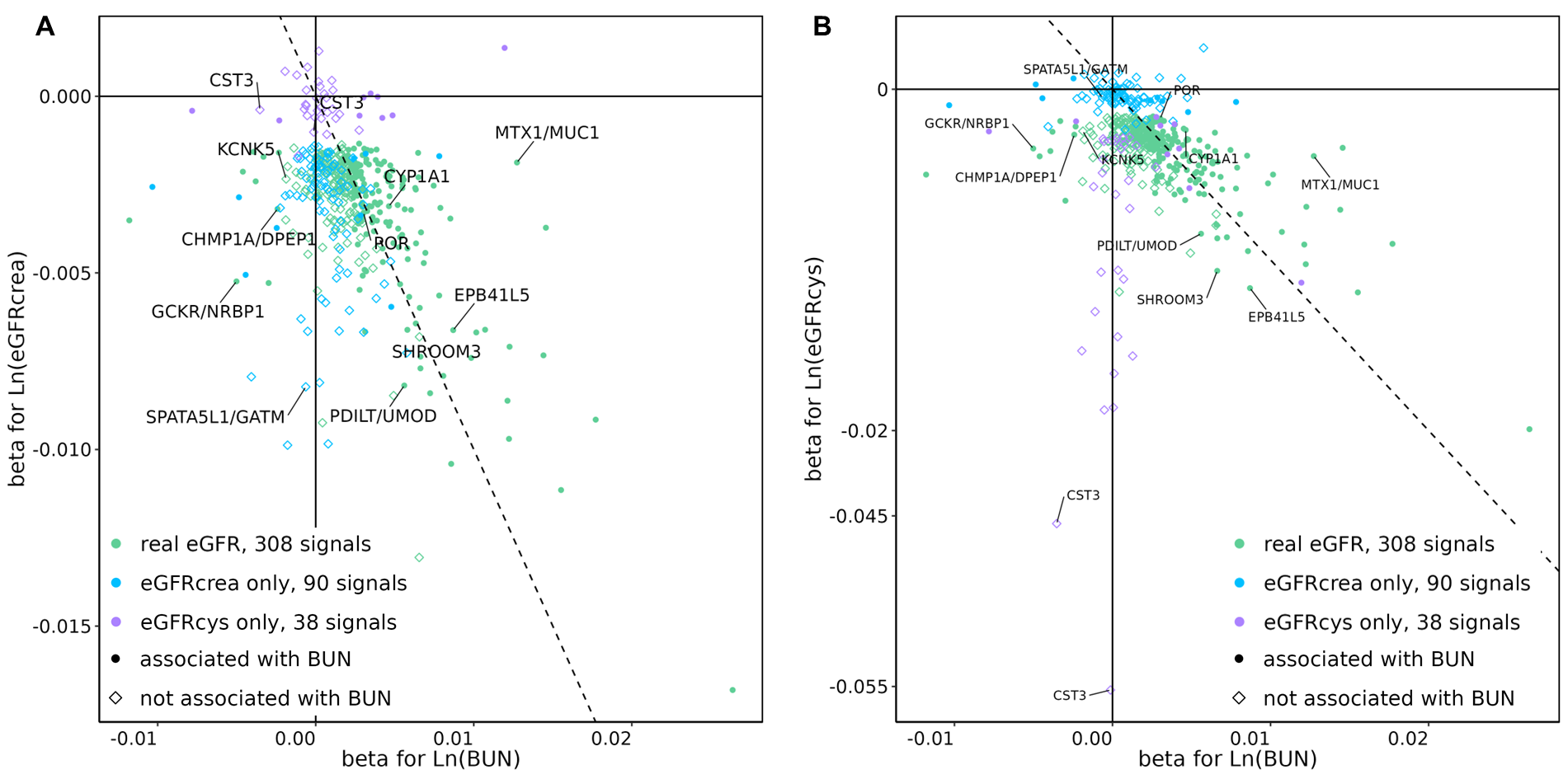

### Supplementary Figure 6: Specific expression per class, gene and cell-type

Mapped genes (**Figure 4**) of the classes “both”, “real eGFR only” and “real UACR only” were queried from the KPMP kidney tissue atlas^1^. The association P-value per cell-type was transformed into the z-score and plotted in the heatmap. The heatmap is separated by class and clustering was performed for genes. FIB: fibroblast; IMM: immune cell; EC: endothelial cell; VSM/P: vascular smooth muscle cell / pericyte POD: podocyte; PEC: parietal epithelial cell; PT: proximal tubule cell; DTL: descending thin limb cell; ATL: ascending thin limb cell; TAL: thick ascending limb cell; DCT: distal convoluted tubule cell; CNT: connecting tubule cell; PC: principal cell; IC: intercalated cell.

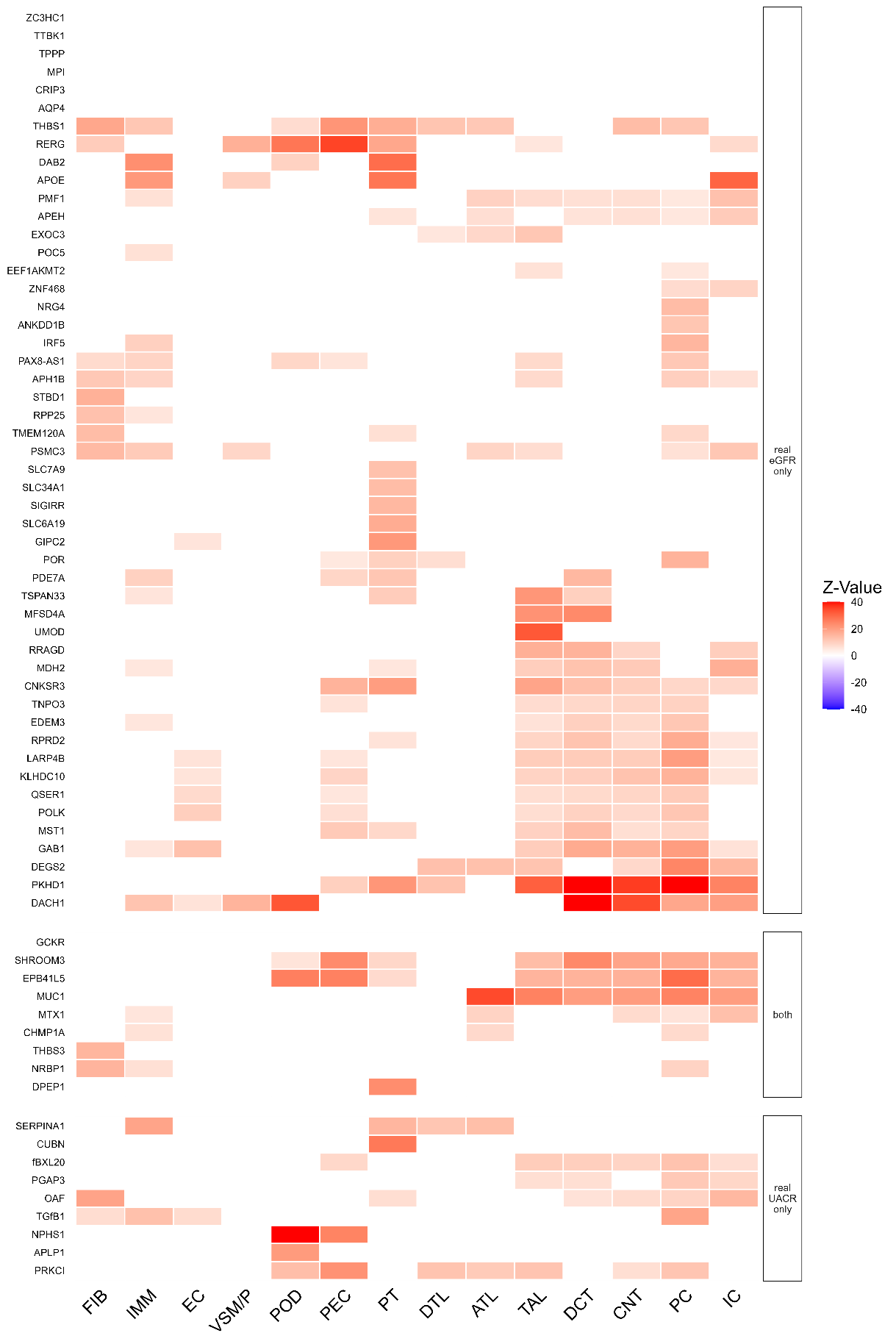

**Supplementary Figure 7: RNA Expression of mapped genes in the “both” group**

Absolut expression was queried and results from the in a combination of healthy and injured cells were screenshotted from <https://shiny.mdc-berlin.de/humAKI/>^2^ (11th July, 2025). RNA expression is depicted as counts per million (CPM) across major kidney cell types. **A**: Absolute RNA expression of five novel mapped genes. **B**: Absolute expression of the *NPHS1* gene. **C**: RNA expression for nine genes mapped in the “both” group (*GTF3C2-AS2* and *LINC02166* were not available). Podo: podocytes; PT: proximal tubules; tL: thin limbs; TAL: thick ascending limb, DCT: distal convoluted tubule; CNT: connecting tubule; CD-PC: collecting duct principal cells; CD-IC-A: collecting duct intercalated cells A; CD-IC-B: collecting duct intercalated cells B; EC: endothelial cells; Leuk: leukocytes; Fibro: fibroblasts.

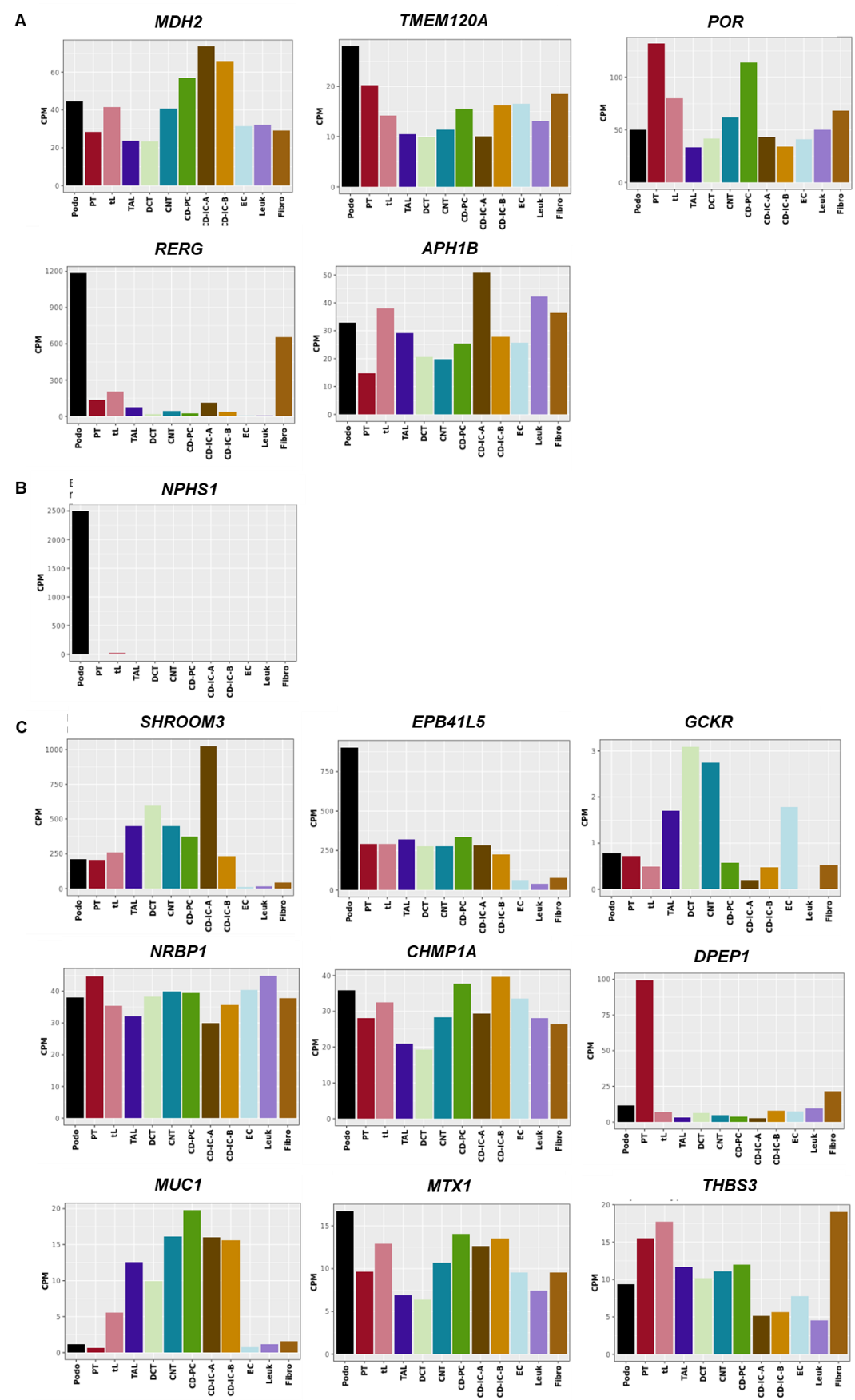

**Supplementary Figure 8: Fluid intake GWAS.**

The fluid intake GWAS was performed on data from UKB (N=436,221). Genome-wide significant (P<5x10^-8^) associated signals were derived using a distance and LD-based criterion (d<500kb, r²>0.01). **A**: The Manhattan plot depicts the result of the fluid intake GWAS. In total, 88 independent signals were found. The top 10 hits were annotated with the nearest gene. **B**: Shown are FUMA-MAGMA tissue enrichment analysis results for the fluid intake GWAS. Tissues marked in red showed significant enrichment of gene expression effects (based on GTEx tissues, false discovery rate, FDR<5%). **C**: Depicted are the genetic effect estimates of fluid intake and Ucrea for the 88 fluid intake index variants. Variants associated with Ucrea are marked by * (P<0.05/88).

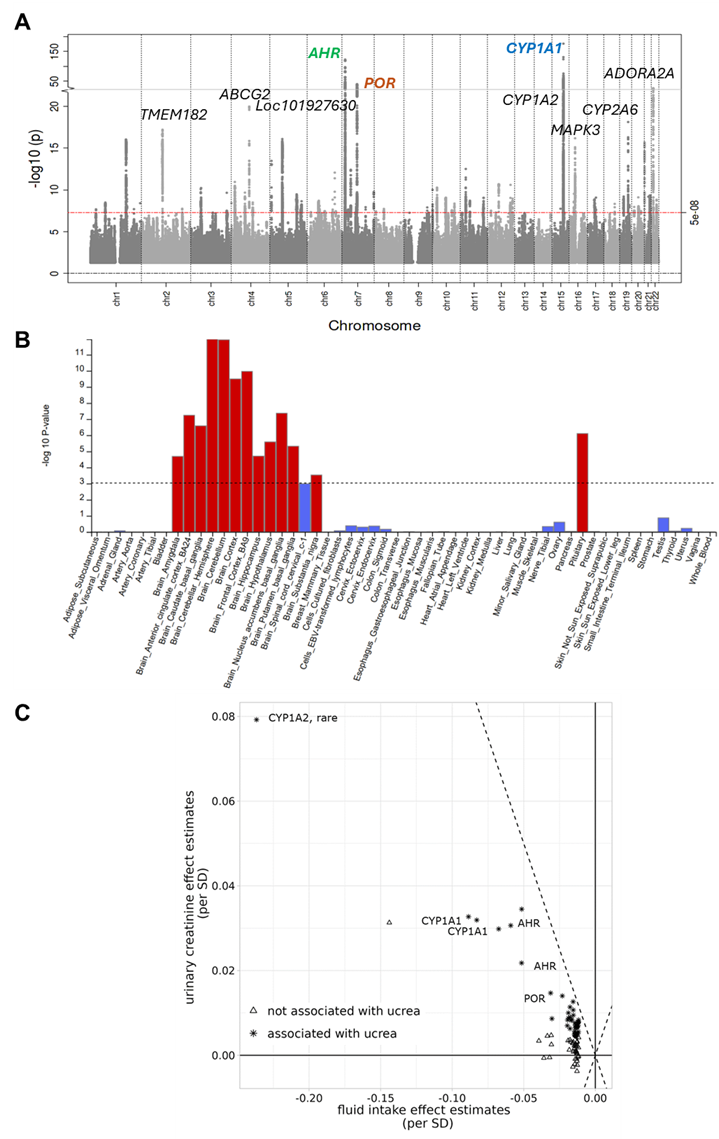

### Supplementary Figure 9: Mediation analyses at the remaining 6 “both” class signals.

For the 11 “both” signals with concordant eGFR and UACR effects, the heatmaps show results from mediator analyses on all combinations eGFR, UACR, MAP and HbA1c for signals not shown in **Figure 8B** (based on unrelated UKB individuals of European ancestry, N~350K, **Methods**). On the diagonal is the total genetic effect on the outcome, off-diagonal are mediated proportions if the effect was significant on the outcome.

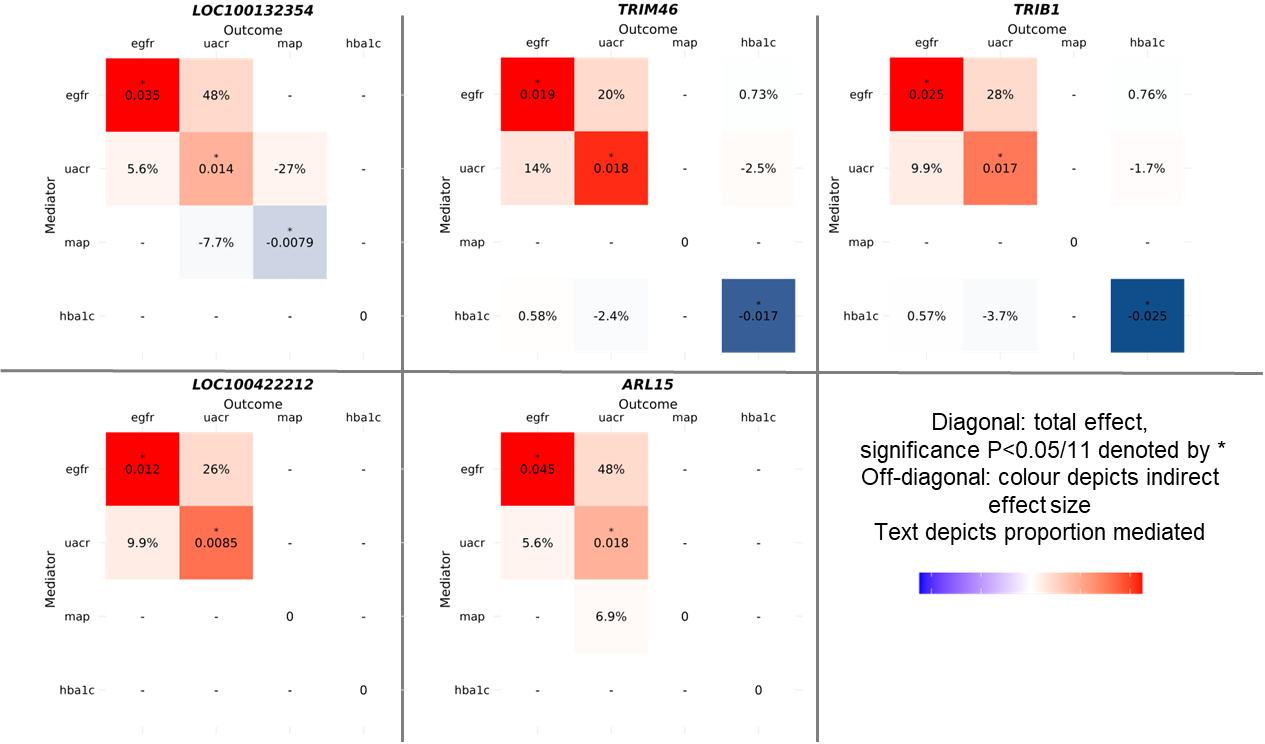

**SUPPLEMENTARY NOTES**

**Supplementary Note 1: Multi-trait GCTA**

Single trait approximate conditional analysis using GCTA^3^ was expanded to be used with multiple traits, using the explained variance of the SNP instead of the association P-value to determine the signal index variant (**Supplementary Figure 4**). Cleaned summary statistics of all four main traits, a list of combined regions and their coordi­nates, and the standard deviation of each main trait was required for this analysis. First, the first genome-wide significant variant is picked as the signal index variant. Next, the explained variance for each variant in the region is calculated for each trait. If the association P-value did not reach P_c,i_<10^-5^, the explained variance for this trait was set to zero before adding it to the explained variances of the other traits. The i-th multi-trait signal index variant, is determined as the variant with the maximum sum of explained variance across all four traits. For each trait, the variants are then conditioned on i identified signal index variants using “cojo-cond”. If any other variant in the region reaches genome-wide significance after conditioning in any of the traits, the explained variance for each variant in the region is calculated for each trait based on the conditioned estimates and set to zero if necessary. Again, the variant with the maximum sum of explained variance across all four traits is chosen as the second signal index variant to be conditioned on in this region. This cycle is repeated I times until no further genome-wide significant variant is found in the region. Then, either the next region is processed or, if all regions have undergone conditioning, a so-called joint approximate conditioning GCTA analysis is performed for each region using “cojo-joint”. This step is calculated for each region in each trait and conditions the multi-trait signals on each other. For example, one region contains three signal index variants rs1, rs2 and rs3. The joint GCTA conditions rs1 on rs2 and rs3, rs2 on rs1 and rs3, and rs3 on rs1 and rs2. This way, fully conditioned estimates for all traits and all multi-trait signal index variants are obtained that allow for proper comparison of effect sizes between traits. GWAS summary statistics from singular secondary and tertiary traits were conditioned on the signal index variants generated by MT-GCTA using joint conditioning to calculate fully conditioned effect estimates for single traits.

**Supplementary Note 2:** **In depth biological characterization of variants at *POR* locus**

At the *POR* region, three genes were mapped by the lead variant rs1057868: the *POR* gene, for which the variant is a missense variant, and two genes, *MDH2* and *TMEM120A*, for which the variant serves as kidney-tissue eQTL. The association of rs1057868 with fluid intake was driven by its strong effect on coffee and tea consumption. This is supported by literature showing association between coffee intake and a 5'UTR *POR* variant rs17685, which is in high-LD with rs1057868^4,5^. *POR* encodes Cytochrome P450 oxidoreductase, essential for CYP function^6^, and its activity can affect CYP enzymes like *CYP1A2*^7^. The correlated SNP rs17685 is associated with increased *POR* activity which could also lead to an increase in *CYP1A2* activity, which has known impact on caffeine preference^4^. In contrast, eQTL-mapped genes mainly affect water resorption. Altered *MDH2,* encoding for the malate dehydrogenase in the citric acid cycle, impacts ATP production^8^ and Na/K-ATPase activity, which affects sodium and water resorption in the tubules^9^. *TMEM120A* (TACAN) interacts with the ion channel Polycystin-2, which is discussed to be involved in mechanosensation and fluid shear stress responses and may promote AQP2 insertion and water resorption via calcium signaling^10,11^. In summary, again, while *POR* could explain the behavioural fluid intake component, *MDH2* and *TMEM120A* may be linked to water handling and kidney function.

**Supplementary Note 3: In depth biological characterization of “both” class signals**

Alleles in the “both”-class affect eGFR and UACR in the same direction. We here discuss potential mechanisms at four of the 11 both class signals:

*SHROOM3*

*SHROOM3* is well-established in kidney biology, implicated in podocyte disruption, glomerular barrier damage, and albuminuria phenotypes in knockout mouse models^12–14^. In our case, the effects of *SHROOM3* are likely not as strong. Still, the high-PIP *SHROOM3* variant might affect podocyte structure, e.g. by making them smaller or the sub-podocyte space bigger. This would lead to bigger slit diaphragms and less ability to handle tensile stress from increased capillary pressure risking podocyte detachment^15^. Additionally, it would increase the area of free filtration, leading to increased filtration of both creatinine and albumin. This would increase the eGFR. The increase in UACR develops after an overload of the reuptake mechanism. This would also explain the effect of eGFR on UACR, since the increase of eGFR is seen before increased urinary albumin is measured. This hypothesis is supported by the notion that *SHROOM3* is differentially expressed in both podocytes and proximal tubular cells (**Supplementary** **Figure 6+7C**). The function of the mapped tubular eQTL is unknown, but it can be hypothesized that it could play a role at this point. Despite the signals association with blood pressure, the mediator effect shows that there is no influence in either direction between blood pressure and eGFR or UACR making a hypothesis using blood pressure to introduce hyperfiltration unlikely^16^.

*EBP41L5*

A similar pattern of association and mediator effects is observed for *EPB41L5* signal. In contrast to *SHROOM3*, the UACR effect is not completely mediated by eGFR and it is expressed not only in podocytes and proximal tubular cells but also in different parts of the Loop of Henle (**Supplementary Figure 6+7C**). *EPB41L5* and its homologues in different model organisms, have been observed to control the assembly of podocyte extracellular matrix and influences maturation of integrin adhesion sites^17–19^. Knockout of this gene results in insufficiently mature adhesion sites, which in turn leads to an impaired transmission of force. This might lead to podocyte detachment and a loss of integrity of the glomerular barrier membrane^17^. Our high-PIP variant maps as a missense variant, meaning that the described effects are possible. These could lead to bigger slit diaphragms and might finally result in an increased eGFR and UACR as described for the *SHROOM3* signal.

*NRBP1*

The high PIP and lead variant in the *GCKR* signal maps as a missense variant to GCKR and as a tubular eQTL to *NRBP1*. *GCKR* is a known Diabetes mellitus (DM) locus, and our variant is also associated with DM. However, mediator analysis showed that there is no influence of HbA1c on either eGFR or UACR, or in the other direction. We were not able to find a hypothesis explaining the influence of the missense *GCKR* variant on eGFR or UACR independently of DM. For the eQTL, it could be explained via influence on renal autoregulation. *NRBP1* has been shown to activate with *WNK* kinases in the kidney. *WNK* signaling activates sodium co-transporters which are needed for sodium re-uptake and pressure control^20,21^. Increased sodium reabsorption leads to less sodium registered at the macula densa, de-activation of the tubule-glomerular feedback, less production of vasoconstrictors and thereby a reduced resistance of the Vas afferens^15,20^. Reduced resistance means that the afferent arteriole becomes wider and the renal blood flow increases. The eGFR and UACR decreasing allele downregulates *NRBP1*, meaning less *WNK* kinase activity and less sodium reabsorption. Less sodium means an activation of the tubule-glomerular feedback with an increase in afferent resistance and a decrease in renal blood flow. This decrease in blood flow results in less filtrate, meaning more creatinine in the serum and less albumin in the urine, resulting in a decreased eGFR and a decreased UACR. Additionally, the variant was queried for eQTL in liver, muscle and pancreas tissue to determine an eventual connection to DM^22^. The variant is not an eQTL in any of the tissues, supporting our hypothesis of pleiotropy with independent effects on Diabetes mellitus and HbA1c and kidney function.

*CHMP1A* and *DPEP1*

The second signal in this subgroup maps to eQTLs for *CHMP1A*, *DPEP1* and *LINC02166*. *CHMP1A* and *DPEP1* have been shown to influence iron export and iron import, respectively, and play a role in iron-mediated cell death^23,24^ (ferroptosis). Iron is essential for ATP synthesis^25^. ATP is in high demand in kidney cells since it is necessary for multiple transporters including ones involved in sodium reabsorption^25,26^. However, both iron overload and iron deficiency can be detrimental^26^. Therefore, iron homeostasis is strictly regulated^26^. Hence, an increase in iron in the cells, driven by *CHMP1A* down- and *DPEP1* upregulation might impact ATP production followed by impaired sodium reabsorption. Less sodium reabsorption might lead as described for *NRBP1* to a lower eGFR. Albumin uptake is also dependent on ATP. The filtrate, however, already contains less albumin, which means less can be reabsorbed and the result is still a lowered UACR since the impairment of reabsorption is covered by the reduced amount of albumin in the filtrate. Long-term iron overload could induce ferroptosis, destroying the cells. This would still lead to a reduced eGFR through the described mechanisms, but the albumin uptake would also be abolished, which would then lead to albuminuria. Since, we have a mostly healthy population these stages of damage have most likely not been reached yet.
